# Information processing speed modulation by electrical brain stimulation in multiple sclerosis: Towards individually-tailored protocols

**DOI:** 10.1101/2024.10.01.24314688

**Authors:** Steffen Riemann, Michel Mittelstädt, Maurice Glatzki, Carlotta Zilges, Clara Wolff, Filip Niemann, Mandy Roheger, Agnes Flöel, Matthias Grothe, Marcus Meinzer

## Abstract

Information processing speed (IPS) is a core cognitive process, highly relevant in everyday-life and the most frequent and disabling cognitive symptom in patients with Relapsing Multiple Sclerosis (pwRMS). By using region-specific focalized transcranial direct current stimulation (tDCS) in healthy individuals and pwRMS, we provide causal evidence for superior parietal lobe (SPL) involvement in IPS and identified a clinical predictor of tDCS-response in pwRMS. The study employed a registered, randomized, sham-tDCS controlled, three-way blinded, cross-over trial and a mixed-factors design with eight arms [between-subjects: group (pwRMS, controls); tDCS-polarity (excitatory, inhibitory); within-subjects: stimulation (active-, sham-tDCS)]. Concurrently with tDCS, participants completed a computerized version of the Symbol-Digit-Modalities-Test (SDMT), the current gold standard for quantifying IPS impairment in pwRMS. Bayesian modeling with generalized linear mixed models provided strong evidence for polarity specific modulation of IPS by SPL-tDCS and a double-dissociation of stimulation response in pwRMS and controls. Healthy individuals showed the canonical pattern of significantly enhanced and reduced processing speed during excitatory or inhibitory tDCS. In pwRMS, a reversed pattern was found and tDCS-response was predicted by baseline SDMT performance; i.e., more or less impaired patients benefited from inhibitory or excitatory tDCS, respectively. Our results provide direct causal evidence for SPL involvement in IPS in health and disease and suggest that the degree of IPS impairment in pwRMS reflects compensatory or dysfunctional neuroplastic processes that can be counteracted in a polarity specific way. Identified standardized transition scores for the effectiveness of excitatory or inhibitory tDCS will inform future individually-tailored stimulation protocols in pwRMS.

**One Sentence Summary:** Polarity-specific modulation of information processing speed in multiple sclerosis.

## INTRODUCTION

Multiple sclerosis (MS) is a chronic inflammatory disease resulting in progressive central nervous system demyelination and neurodegeneration.^1^ The relapsing form of MS (RMS) is the leading cause of non-traumatic neurologic disability in early adulthood worldwide and manifests as recurrent episodes of inflammation, followed by variable degrees of remission and progressive neurological dysfunction.^2^ Patients may suffer from physical and psychological symptoms, and up to 65% of patients present with cognitive impairment.^3^ Cognitive impairments predicts later disability, and is one of the main reasons for reduced work productivity and associated with reduced quality-of-life.^4–8^

The most prevalent and disabling cognitive symptom in people with RMS (pwRMS) is reduced information processing speed (IPS).^3,9^ IPS is a key cognitive process that comprises the ability to identify, discriminate, integrate, and make decisions about incoming information, and to swiftly respond at the behavioral level. According to the tri-factor model of IPS, sensory, cognitive and motor speed components can be distinguished that are subserved by a brain network comprised of sensory, fronto-parietal and cerebellar regions.^10^ It is estimated that 27-51% of pwRMS patients suffer from IPS impairment, which has been linked to disease progression, but can also be the only symptom of a relapse.^3^

Drug treatment of the inflammatory neuropathology cannot directly target impaired IPS and there are currently no approved medications for treating cognitive symptoms in pwRMS.^3,11^ This highlights the clinical relevance of exploring novel and evidence-based treatments that directly modulate activity and plasticity in the neural networks subserving IPS in pwRMS.^9^ A promising approach to achieve this is non-invasive transcranial direct current stimulation (tDCS), that administers weak electrical currents via scalp attached electrodes to the brain, to modulate neural excitability and plasticity in the human brain.^12^ TDCS has an excellent safety profile and has successfully been used to improve cognitive functions in healthy individuals and patients with neuropsychiatric disorders.^13–19^

To date, the vast majority of tDCS studies in pwRMS have targeted motor symptoms or fatigue and only two studies specifically investigated tDCS effects on IPS.^20,21^ These studies administered multisession tDCS to the prefrontal cortex, either as stand-alone treatment, or in combination with cognitive training.^22,23^ Only the latter reported improved IPS immediately after the treatment relative to a control group, but no between-group effects six months later. These mixed effects of tDCS mirror those reported for other symptoms in pwRMS.^20,21^

Moreover, animal and human studies have demonstrated that behavioral and neural effects of multisession tDCS can be maintained for weeks or even months after the end of the intervention period.^24–27^ In this context, it is worth noting that reorganization of the functional brain networks supporting cognition in pwRMS is poorly understood and upregulation of fronto-parietal activity in pwRMS may reflect compensatory or dysfunctional neuroplasticity, depending on disease progression.^28–33^ This makes it difficult to predict the neural effects of specific tDCS montages and unintended long-term stimulation effects in multisession contexts bear the risk for inducing maladaptive neuroplasticity. Hence, there is currently an urgent need for conducting carefully designed proof-of-concept tDCS studies (1) that investigate the potential of specific tDCS interventions to enhance IPS and (2) to identify clinically relevant predictors of stimulation response, prior to conducting time- and cost-intensive interventional trials, while minimizing the risk for the patients.

This was accomplished in the present study, by implementing a balanced and randomized crossover trial that involved two stimulation sessions, either active or placebo (sham) tDCS. A single active tDCS session was chosen, because effects are completely reversible, thereby minimizing the risk for potential negative long-term effects.^13^ During each session, pwRMS and age- and sex matched healthy individuals completed a computerized version of the Symbol Digit Modalities Test (SDMT), which is the current gold standard to determine the degree of processing speed impairment in pwRMS.^34^ TDCS was administered bilaterally to the superior parietal lobe (SPL), based on previous functional and diffusion imaging studies, suggesting specific involvement of this region in the speed component of IPS.^35,36^ A focalized tDCS set-up was used to constrain the current delivery to the target regions, which has been shown to result in regionally- and task-specific neural modulation.^37–41^ By investigating polarity specific (i.e., anodal-excitatory-cathodal-inhibitory; AeCi) effects of focal tDCS, we aimed to establish causal involvement of the SPL in IPS. Inclusion of a matched healthy control group served to determine the degree of IPS impairment in pwRMS (i.e., during sham tDCS) and to investigate the question if the expected AeCi pattern of active tDCS in healthy controls can be replicated in the patient groups. Based on functional imaging studies suggesting that fronto-parietal activity may reflect compensatory or dysfunctional processes depending on disease progression, we also explored if tDCS-response in the patients can be predicted by the degree of IPS impairment.^28–33^

## RESULTS

This prospective, randomized, three-way blinded (i.e., participants, staff conducting the experiment and evaluating outcomes), sham-tDCS controlled, cross-over trial was conducted in a mixed-factors design with eight arms, representing the between-subjects factors group (pwRMS, healthy controls; N=32/group) and tDCS polarity (anodal, cathodal), and the within-subjects factor stimulation (active, sham tDCS). All participants completed a baseline phase, including comprehensive neuropsychological assessments (**Table 1**), followed by the experimental cross-over phase (**Figure 1A**). During each phase, participants completed the computerized SDMT (**Figure 1B+2**) and received either active or sham SPL-tDCS (**Figure 3**). Response latency was defined as primary outcome (Clintrials.gov: NCT04667221); accuracy data was also analyzed to confirm compliance with task instructions to answer as fast and accurate as possible. Statistical analyses were conducted in a Bayesian framework and data were analyzed using generalized linear mixed models (GLMMs) with lognormal and binomial distribution for response latency and accuracy data. Three models were fitted for each analyses to identify the best fitting model for our data: (1) An intercept only model; (2) a covariates model to correct for potential session effects, motor slowing (via the Trail Making Test A, TMT-A) and baseline performance on the paper-pencil SDMT;^42^ (3) A full model comprising the covariates specified above, and variables of interest (i.e., tDCS polarity [anodal, cathodal], group [pwRMS, controls], and stimulation type [active, sham]), as well as their interactions. Models included a group-level subject intercept (random intercept) to correct for individual performance levels. To establish the model that best explained our data, models were compared using the widely applicable information criterion (WAIC), which is a fully Bayesian extension of the Akaike information criterion (AIC), and the estimated log pointwise predictive density of the WAIC (ELPD WAIC; see Methods for details).^43^ Bayesian hypothesis testing used evidence ratios, which considers the evidence for and against specific hypotheses; i.e., possible changes in performance due to repeated task exposure would be investigated with the following formula: 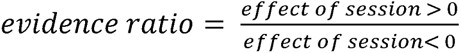. For a detailed description of the models and interpretation of evidence ratios see Methods & Materials.

**Figure 1.**
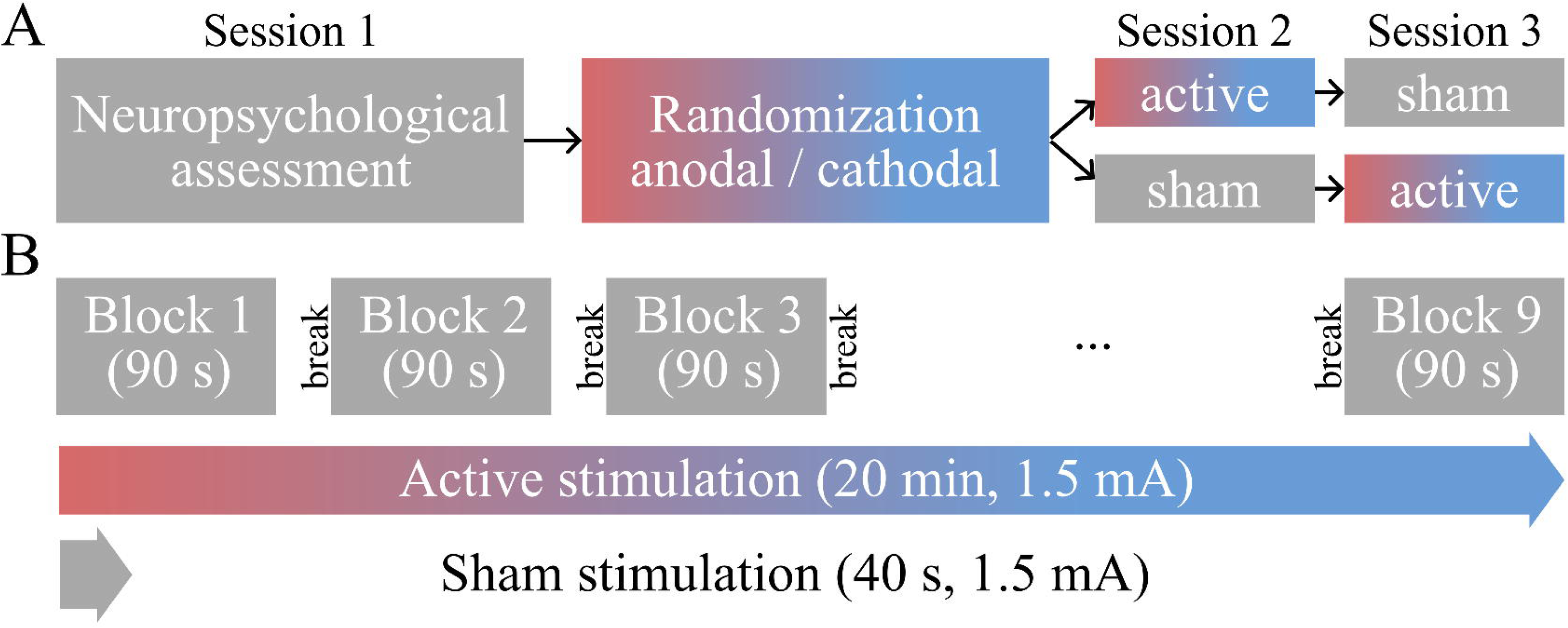
Study overview. **(A)** The study comprised three sessions: A neuropsychological baseline assessment, followed by two experimental sessions (either active or sham tDCS); stimulation order was counterbalanced in each participant group. **(B)** Procedure of the experimental sessions: Nine blocks of the modified SDMT were completed with either active (anodal, cathodal) or sham stimulation. Breaks in between blocks were self-paced, but could not exceed 60 sec. tDCS=transcranial direct current stimulation, SDMT=Symbol-Digit-Modalities Test.

**Figure 2.**
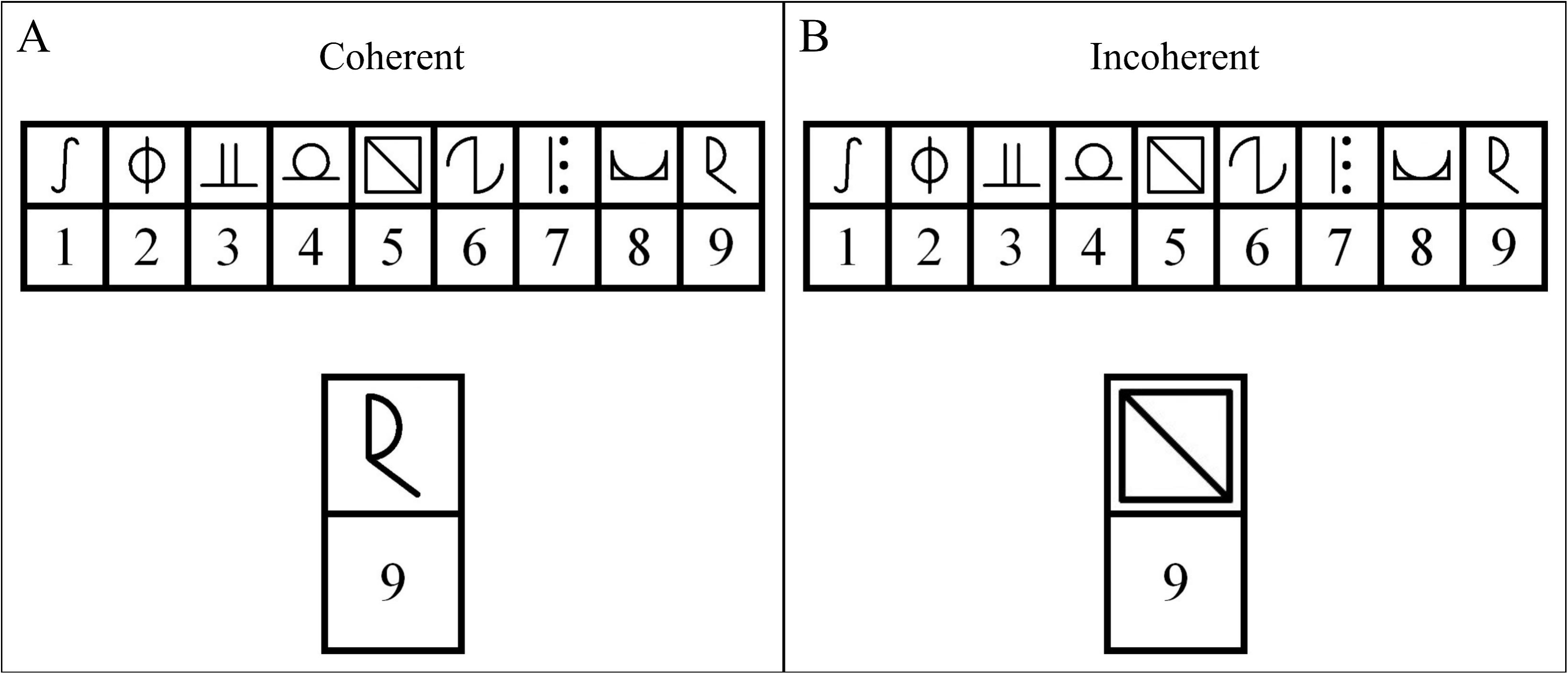
Experimental task: Examples of **(A)** coherent and **(B)** incoherent trials of the modified SDMT. At the top of the image the legend of the current block is displayed, which indicates correct symbol-digit combinations. At the bottom, a ‘probe’ combination is shown and participants had to indicate coherent or incoherent combinations by button-press. SDMT=Symbol-Digit-Modalities Test.

**Figure 3.**
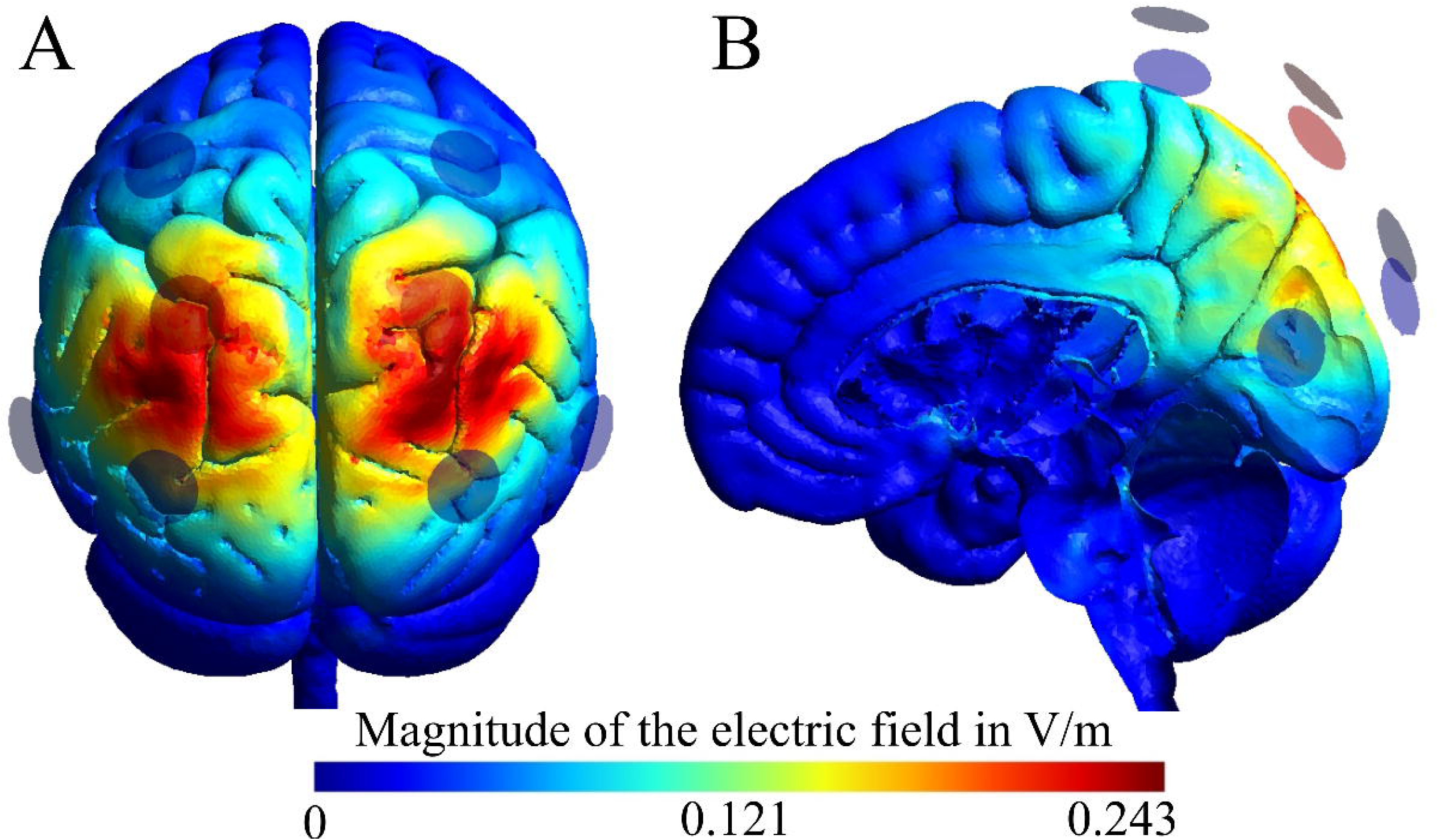
TDCS set-up and current flow simulations for bilateral SPL-tDCS montage. The bilateral montage was comprised of a focalized 3×1 set-up, with two central electrodes (red) and three surround electrodes (blue) that constrain the current flow to the target regions. The set-up was chosen during the planning stage of the project based on current flow simulations using standard SimNIBS parameters and a MNI152 standard brain^64,65^ **(A)** Shows peak current intensity in the target region (bilateral superior parietal lobe); **(B)** Medial view illustrates current intensities to deeper brain regions. Note: Current flow patterns are shown for the excitatory “anodal” tDCS condition; cathodal stimulation has a reversed polarity, but identical current distribution. SPL=superior parietal lobe, tDCS=transcranial direct current stimulation, V/m=Volt/Meter.

**Table 1.**
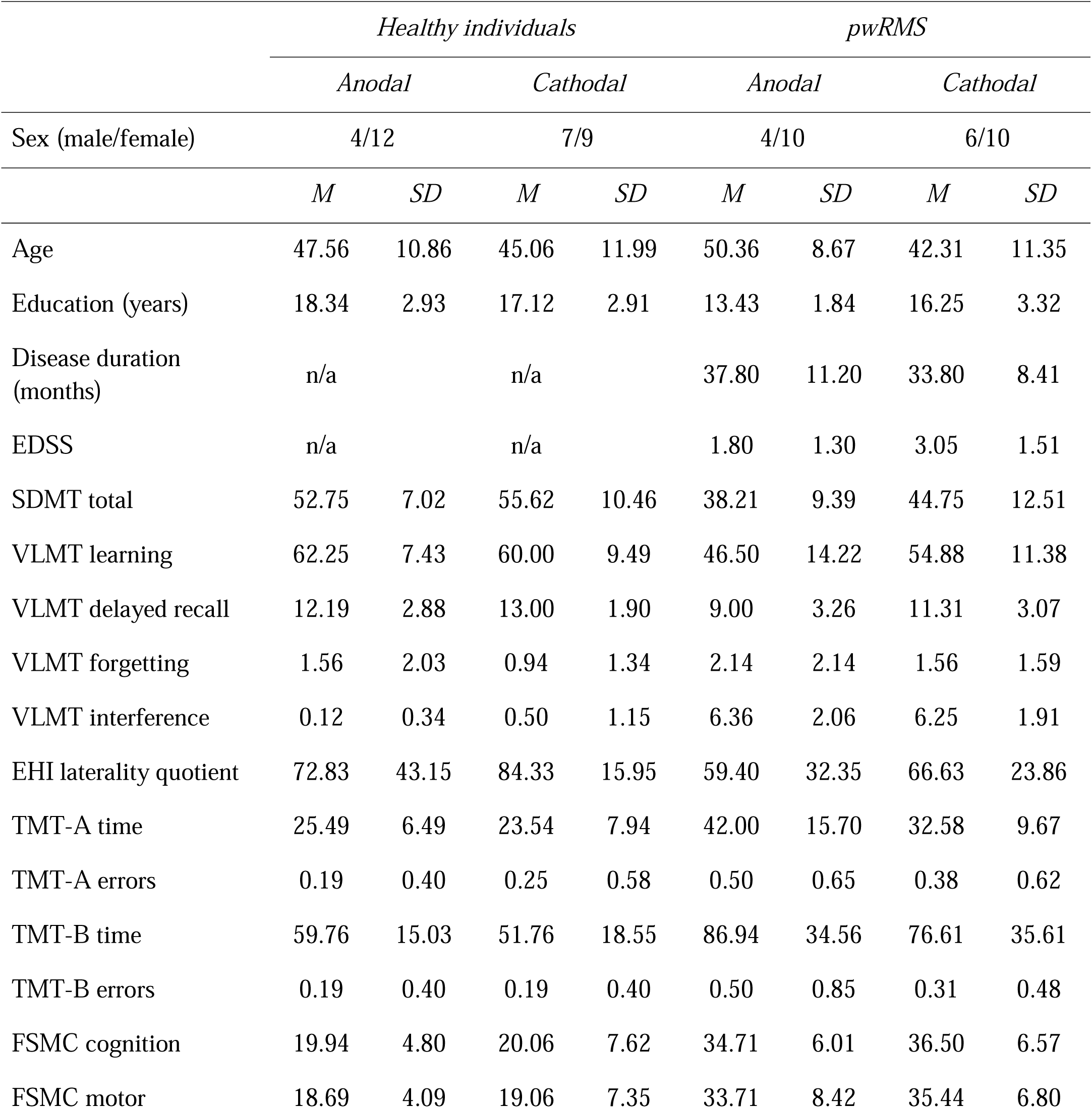
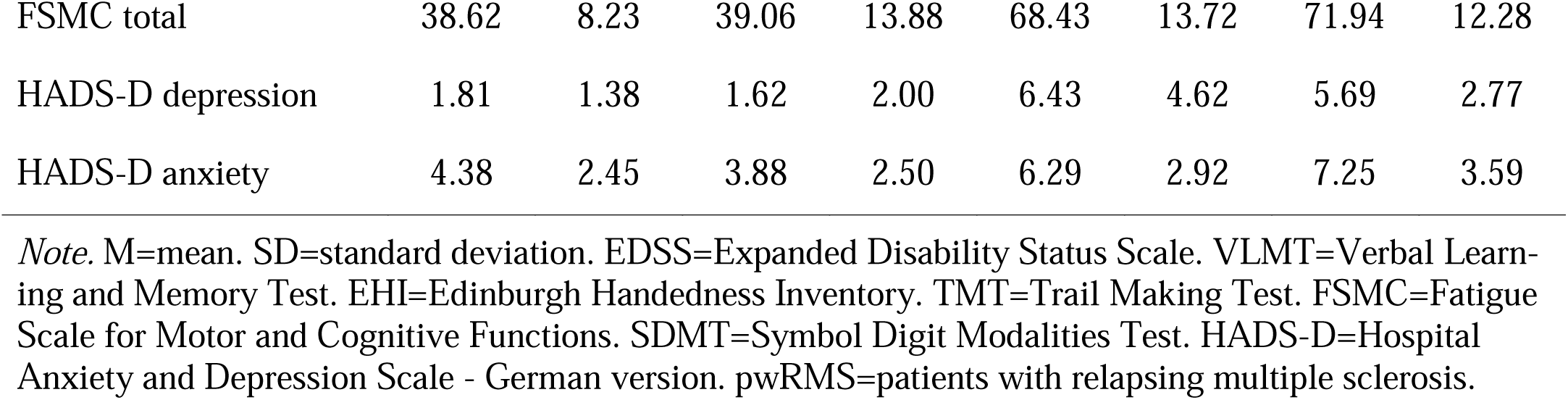
Demographic and clinical characteristics of patients and healthy controls and details of the neuropsychological assessment (Note: raw scores are reported; all healthy participants scored within agecorrected norms)

### Response accuracy

Response accuracy during the experimental SDMT task was near ceiling level across groups (*M*=98.23%, *SD*=13.2%); group means are shown in **Table S1**, indicating compliance to the instructions to answer as fast as possible without making mistakes. WAIC revealed that the intercept only model performed best (**Table S2**), suggesting that differences in accuracy levels were not explained by the factor group or the stimulation conditions (**Table S3**).

### TDCS effects on information processing speed

This analysis specifically addressed effects of the respective tDCS conditions on IPS in healthy individuals and pwRMS. Assessment of the response latency models showed that the full model including covariates and the effects of interest performed best, indicated by lower ELPD WAIC and WAIC values (**Table S4**). The full model for response latency provides strong evidence for a three-way interaction between the effects of interest: Group × stimulation type × polarity (*β*=- 0.10, 95%CI=[-0.11, -0.08], *evidence ratio=*∞; **Figure 4**). In the control group, post-hoc tests showed that anodal stimulation decreased reaction times (*β=*-0.03, 95%CI=[-0.03, -0.02], *evidence ratio*=∞) and cathodal stimulation increased reaction times (*β*=0.04, 95%CI=[0.04, 0.05], *evidence ratio*=∞) compared to their respective sham conditions. However, in the patients, cathodal stimulation decreased response latencies (*β*=-0.02, 95%CI=[-0.02, -0.01], *evidence ratio=*4,443.44), while anodal stimulation increased response latencies (*β*=0.01, 95%CI=[0.01, 0.02], *evidence ratio*=929.23) relative to sham tDCS. Hence, we demonstrate a double dissociation for effects of anodal vs. cathodal tDCS in pwRMS and healthy controls (**Table S5**).

**Figure 4.**
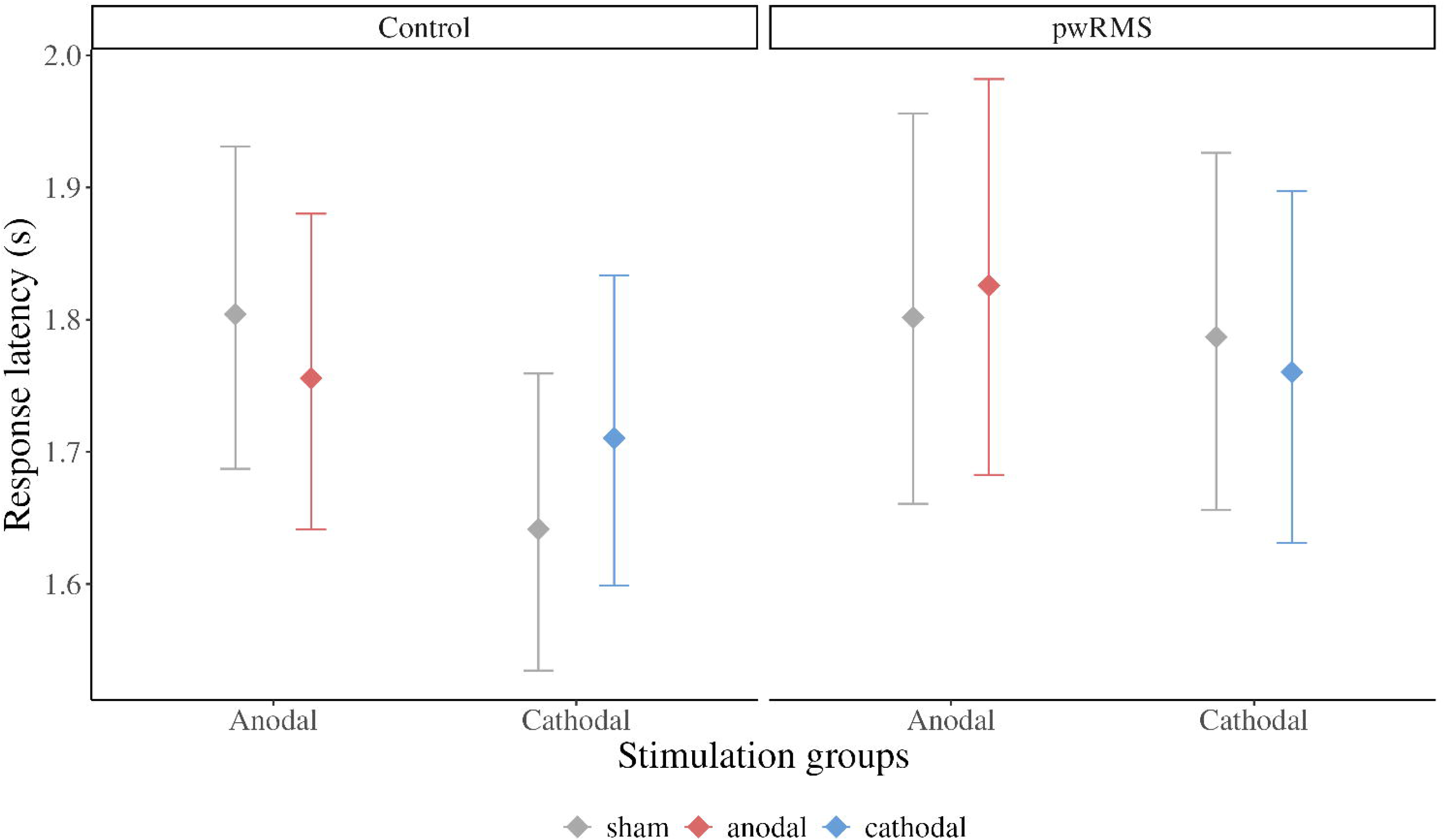
Response latency effects. Conditional effects plot for response latency effects of anodal and cathodal stimulation in both subject groups, controlled for session effects, motor slowing (via TMT-A), and the number of correct responses in the paper-pencil SDMT. Compared to sham stimulation, healthy controls exhibited shorter response latencies during anodal stimulation and increased response latencies during cathodal stimulation. Across the groups, this effect was reversed in pwRMS, i.e., response latencies were shorter during cathodal stimulation and longer during anodal stimulation (relative to sham stimulation). pwRMS=patients with relapsing multiple sclerosis.

Additionally, there was strong evidence that the covariates included in our model influenced latency in the experimental SDMT. Participants were faster during the second session suggesting a learning effect (*β*=-0.12, 95%CI=[-0.12,-0.11], *evidence ratio=*∞). Longer response latencies in the TMT-A were positively correlated with longer response times in the experimental SDMT (*β*=0.06, 95%CI=[0.01, 0.10], *evidence ratio*=129.72), i.e., the term corrected for motor slowing. Additionally, an increased number of correct responses in the baseline paper-pencil SDMT, i.e., an indirect measure of shorter response latencies during the baseline SDMT, was associated with shorter response latencies in the modified SDMT (*β*=-0.17, 95%CI=[-0.21, -0.12], *evidence ratio=*∞). Therefore, this covariate corrected for group differences in the baseline SDMT performance and the results provide strong evidence for a correspondence between the clinical, and experimental versions of the SDMT (i.e., construct validity).

### Baseline IPS performance predicts tDCS response

To investigate if baseline performance on the clinical version of the SDMT predicted tDCS response, the full reaction time model was extended: Baseline performance on the paper pencil SDMT, group, stimulation polarity and stimulation type, as well as their interactions were added as population level effects. Additionally, we corrected for TMT-A performance and the session effect, i.e., performance increase due to repeated exposure to the task. Subjects were added as group-level intercepts. To ensure the validity of this analysis and enhance clinical relevance of potential outcomes, we considered two different predictors: (1) study specific raw-scores of the baseline SDMT in our sample, (2) age- and education corrected norms of the SDMT.

Overall, results were consistent for the study specific raw-scores of baseline SDMT performance and the analysis that used age-corrected norms (with better model fit for study specific data over the previous full model; ELPD difference=-255.27; SE difference=25.12; |-255.27| > 49.23).^34^

A four-way interaction between group, polarity, stimulation type, and baseline SDMT score was confirmed for the study-specific and norm-based analyses (study specific: *β*=0.06, 95%CI=[0.04, 0.08], *evidence ratio=*∞; age-corrected norms: *β*=0.06, 95%CI=[0.04, 0.08], *evidence ratio=*∞; **Figure 5**). This four-way interaction has to be interpreted with caution because the group factor and baseline SDMT scores are related, i.e., patients generally have lower SDMT scores than healthy controls. The latter is illustrated in the upper panels of **Figure 5B** (i.e., lack of z-scores lower than -1 in healthy controls). Nonetheless, healthy controls with higher baseline SDMT scores showed a stronger canonical AeCi response to tDCS, i.e., anodal stimulation reduced response latency and cathodal stimulation increased response latency, relative to sham stimulation. Because none of the healthy controls had z-scores lower than -1, only the canonical response was observed.

**Figure 5.**
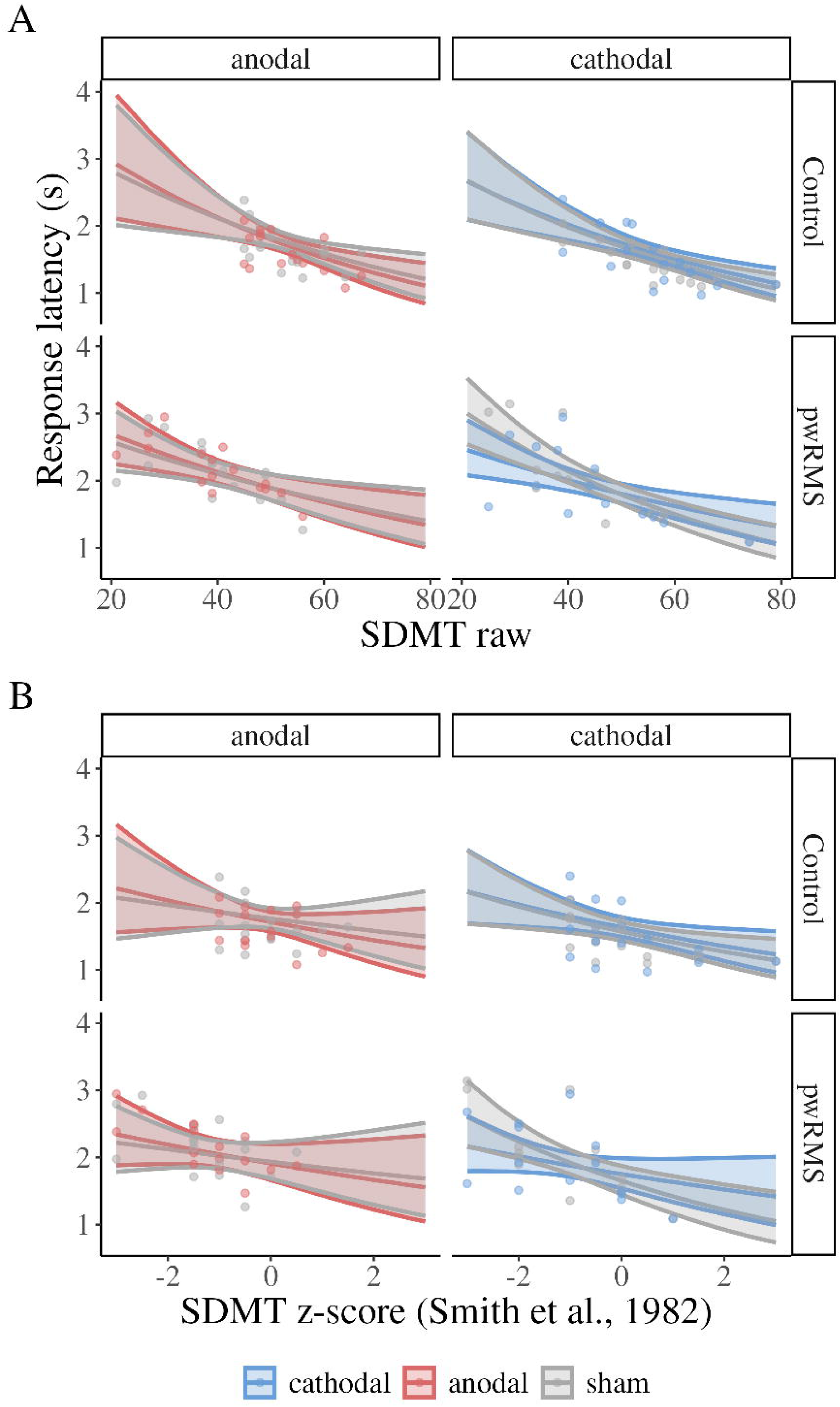
Conditional effects plot. Illustrates the three-way interactions for the association between response latency during either placebo (sham) and anodal or cathodal tDCS. **(A)** Study specific distribution of raw scores of the baseline SDMT. **(B)** Age corrected SDMT norms.^34^ Controls did not reach z-scores smaller -1 indicating that the patient group drives the reversal of the AeCi response. The x-axis shows the raw or z-scores of the baseline. Note the raw SDMT scores were standardized during model computation, but rescaled for plotting (M=48.15, SD=11.93). The y-axis shows mean response latency during sham or active (anodal; cathodal) tDCS conditions. Ribbons represent a 95% credible interval. Dots represent mean response latency of individual participants across experimental blocks and baseline SDMT scores of the participants.

However, the four-way interaction also indicates that the moderating effect of baseline SDMT was larger in pwRMS. In addition, since we included patients with MS that had z-scores lower than -1, we observed a reversal of the canonical AeCi response, i.e., cathodal reduced response latency and anodal increased response latency (**Tables S6+7)**. Notably, for standardized scores, the transition from beneficial to non-beneficial effects of anodal (z <-.58) or cathodal (z >-.70) was consistent across the patient groups.

We additionally repeated the analyses and only included the patient-subgroups, to investigate if the control group drove the interaction effect. Here, we found a three-way interaction of stimulation polarity, stimulation type and baseline SDMT score, indicating that this was not the case (**Tables S8+9**).

### Adverse effects, mood and affect and blinding efficacy

Overall, participants tolerated the stimulation well, there was no evidence that potential changes in mood and affect biased the behavioral tDCS effects and participant blinding was successful. Details of the statistical models investigating adverse effects, mood and affect and blinding efficacy are described in the Methods and Materials section.

Self-reported adverse effects were mainly rated as “none” (81.73%) or “mild” (14.05%). “Moderate” and “strong” adverse effects were reported in 0.6-3.63% of participants (**Table 2**). Comparison of adverse effects models favored the simple model assuming that the strength of adverse effects is similar in all experimental groups since the absolute ELPD difference was smaller than the standard error multiplied by 1.96 (ELPD difference=-2.4; SE difference=2.9; |-2.4| < 5.68). A post-hoc test showed a beta value for stimulation around 0, and a small increase of evidence from the prior model (*β*=0.03, 95%CI=[-0.18, 0.23], *evidence ratio*=9.44), suggesting that adverse effects were comparable between active and sham conditions (**Table S10**).

**Table 2.** Mean adverse effects score, when assigning 0-3 to none, mild, moderate, and strong adverse effects.

| Sensation | Control |  |  |  |  |  |  |  | pwRMS |  |  |  |  |  |  |  |
| --- | --- | --- | --- | --- | --- | --- | --- | --- | --- | --- | --- | --- | --- | --- | --- | --- |
|  | anodal |  |  |  | cathodal |  |  |  | anodal |  |  |  | cathodal |  |  |  |
|  | sham |  | active |  | sham |  | active |  | sham |  | active |  | sham |  | active |  |
|  | *M* | *SD* | *M* | *SD* | *M* | *SD* | *M* | *SD* | *M* | *SD* | *M* | *SD* | *M* | *SD* | *M* | *SD* |
| heat | 0.25 | 0.45 | 0.50 | 0.63 | 0.19 | 0.40 | 0.12 | 0.50 | 0.14 | 0.53 | 0.06 | 0.25 | 0.07 | 0.27 | 0.21 | 0.58 |
| itching | 0.25 | 0.45 | 0.31 | 0.48 | 0.56 | 0.73 | 0.25 | 0.45 | 0.00 | 0.00 | 0.31 | 0.60 | 0.14 | 0.53 | 0.21 | 0.58 |
| burning | 0.19 | 0.40 | 0.50 | 0.63 | 0.69 | 0.70 | 0.38 | 0.50 | 0.00 | 0.00 | 0.25 | 0.58 | 0.07 | 0.27 | 0.21 | 0.58 |
| pain | 0.19 | 0.54 | 0.19 | 0.40 | 0.19 | 0.54 | 0.06 | 0.25 | 0.07 | 0.27 | 0.06 | 0.25 | 0.21 | 0.43 | 0.29 | 0.61 |
| metallic taste | 0.00 | 0.00 | 0.00 | 0.00 | 0.00 | 0.00 | 0.00 | 0.00 | 0.00 | 0.00 | 0.00 | 0.00 | 0.00 | 0.00 | 0.00 | 0.00 |
| fatigue | 0.44 | 0.81 | 0.19 | 0.54 | 0.31 | 0.48 | 0.25 | 0.45 | 0.43 | 0.85 | 0.38 | 0.62 | 0.43 | 0.76 | 0.43 | 0.94 |
| other | 0.19 | 0.40 | 0.06 | 0.25 | 0.19 | 0.40 | 0.00 | 0.00 | 0.43 | 0.65 | 0.75 | 1.00 | 0.64 | 1.01 | 0.64 | 0.84 |

*Note.* M=mean. SD=standard deviation. pwRMS=patients with relapsing MS.

Potential effects of tDCS on (positive and negative) mood and affect were assessed before and after each stimulation session using the Positive and Negative Affect Schedule (PANAS).^44^ Comparison of the PANAS models favored the simpler model, since the full model only yielded marginally better model criteria (ELPD difference=-0.5; SE difference=2.5; |-0.5| < 4.9). This suggests that PANAS change scores (i.e., preto post active vs. sham tDCS) were comparable in all experimental groups. Post-hoc tests showed weak evidence that the reported affect changes from pre-to post-stimulation is the same during sham and active stimulation in all groups (*β*=0.21, 95%CI=[-1.11, 1.55], *evidence ratio*=1.45). The full model and summary statistics (groups, valence) are reported in **Tables S11+12**.

Assessment of blinding showed that many participants reported to either “not know”, when they were stimulated (41.94%) or reported to be stimulated during the second session (37.10%), the latter suggesting a recency-effect. Model comparisons between the three models showed that the full model performed slightly better than the simple model (ELPD difference=-3.3; SE difference=1.4; |-3.3| > 2.81). The full model, however, did not show a clear pattern for the response behavior of participants. Most pronounced evidence was found for a different response pattern in pwRMS. They were more likely to answer that they did not know when they were stimulated and less likely to answer that they think they were stimulated during the second session (*β*=-0.91, 95%CI=[-2.12, 0.28], *evidence ratio*=13.32; **Figure S1A**). Similarly, people receiving cathodal stimulation during the second session were more likely to report that they did not know when they were stimulated and less likely to report that they thought they were stimulated during the second session (*β*=-1.09, 95%CI=[-2.52, 0.36], *evidence ratio*=13.00; **Figure S1B**). However, the posterior distributions for all predictors were relatively wide, which does not allow for a clear interpretation of effects. Hence, it is concluded that blinding of participants was successful. For details see **Table 3** and **Table S13**.

**Table 3.** Counts for responses to the question: “When do you think you were stimulated?“.

| Group | Stimulation polarity | Active Session | Do not know | Session 1 | Session 2 |
| --- | --- | --- | --- | --- | --- |
| Control | anodal | Session 1 | 2 | 3 | 3 |
|  |  | Session 2 | 3 | 1 | 4 |
|  | cathodal | Session 1 | 0 | 1 | 7 |
|  |  | Session 2 | 4 | 4 | 0 |
| pwRMS | anodal | Session 1 | 3 | 3 | 2 |
|  |  | Session 2 | 5 | 0 | 3 |
|  | cathodal | Session 1 | 5 | 0 | 1 |
|  |  | Session 2 | 4 | 1 | 3 |

*Note.* Grey background indicates the session, in which stimulation was applied.

## DISCUSSION

This study presents strong evidence for causal involvement of bilateral SPL in the speed component of IPS in health and disease. Moreover, by identifying a clinically relevant participant-dependent predictor of stimulation response in pwPMS (i.e., baseline clinical SDMT performance), we provide a framework for future individualized neurostimulation treatments of impaired IPS in this population. Our main findings can be summarized as follows: (1) Relative to sham-tDCS, healthy individuals showed the expected polarity specific pattern of increased (inhibitory tDCS) or decreased (excitatory tDCS) response latency during the experimental SDMT, which provides strong causal evidence for involvement of bilateral SPL in healthy individuals. (2) At the group level, the opposite pattern was found in the patient groups and there was strong evidence for a double dissociation of tDCS effects between pwRMS and age- and sex matched healthy controls. This emphasizes that stimulation effects observed in healthy individuals may not necessarily translate in a one-by-one fashion to clinical populations with structural and functional brain reorganization. (3) However, our results also demonstrate that the degree of baseline IPS impairment in the patient groups mediated stimulation response in a polarity specific way. While more impaired patients responded favorably to cathodal tDCS (transition at z=-0.70), less impaired patients improved more when receiving anodal tDCS (transition z=-.58). This result is in line with functional imaging studies in pwRMS (for review see^28^), that have suggested compensatory upregulation of metabolic brain activity in task-relevant brain networks in patients with less pronounced motor or cognitive impairment. In more impaired patients, enhanced activity may indicate dysfunctional processes, involving breakdown of functional network communication or disinhibition. Notably, the predictive value of baseline SDMT impairment for stimulation response in pwRMS was confirmed for both the study specific distribution of baseline SDMT values and standard scores, which will be particularly valuable to inform clinical tDCS treatment decisions in the future.^34^ (4) Finally, the rigorous design of our study and the high level of experimental control ensured that our results cannot be explained by unspecific factors (e.g., unblinding of participants or staff, tDCS effects on mood or affect, order effects in the cross-over phase). In addition, adverse effects were minimal and comparable between groups and stimulation conditions, which highlights safety, tolerability and feasibility of our focalized tDCS approach.

Over the last two decades, tDCS has been studied extensively for its potential to improve human brain function. The majority of this research has focused on motor functions (Stagg & Nitsche 2018 JECT), which has also been a major focus in pw(R)MS.^20,21^ However, there is increased evidence that higher cognitive functions, that are known to be supported by large-scale neural networks^45^ can be improved by tDCS. Notably, stimulation administered to key nodes of task-relevant cognitive networks has shown promise to enhance performance and brain function in healthy individuals and neuropsychiatric diseases.^14,46^, but also revealed substantial variability of tDCS response within and across studies. The underlying sources of this variability are thought to be multifactorial and can broadly be described as participant-dependent (e.g., skull and brain anatomy or functional network organization) and stimulation-dependent factors (e.g., tDCS timing, duration, intensity or focality), as well as their interactions.^12,47^ Moreover, this research has highlighted that structural and functional brain organization due to normal aging or in patients with neuropsychiatric diseases can alter stimulation response relative to healthy young or age-matched reference populations, respectively.^15,46^ This emphasizes the necessity to conduct carefully designed proof-of-concept studies to investigate the effects of specific tDCS protocols in clinical populations and to identify predictors of stimulation response, prior to implementation in time- and cost-intensive clinical trials. This is particularly relevant in pwRMS, where less than a handful of studies have investigated potential tDCS effects on cognition and yielded mixed results.^20,21^

Consequently, the present study implemented a highly controlled, prospective, randomized, triple-blinded, balanced cross-over trial, that involved assessment of polarity effects and employed a focalized tDCS set-up, aimed at confirming the causal role of the SPL in IPS in pwRMS and matched healthy controls. The relevance of the stimulation target was informed by imaging studies in pwRMS that suggested involvement of bilateral SPL in the speed component of IPS, fMRI adaptations of the SDMT resembling the task used in the present study, and also evidence from lesion studies demonstrating specific involvement of SPL in the manipulation and re-arrangement of information in working memory, which is highly relevant for the SDMT.^35,36,48,49^ Moreover, the adapted experimental paradigm was designed to minimize dependence on fine-motor skills compared to the clinically used paper-and-pencil version and allowed increasing the number of trials to enhance statistical power to detect potential tDCS effects.^34^ Our results also provide strong evidence for a correspondence between the novel computerized and paper-pencil versions of the SDMT, which highlights construct validity of the experimental paradigm employed in this study. Using this task and overall research design, we were able to confirm causal involvement of the SPL in SDMT performance in healthy controls and pwRMS. In addition, we identified a clinically relevant predictor of stimulation response in the patient groups (clinical SDMT impairment), that may be valuable to guide future tDCS treatment studies.

The canonical anodal-excitatory, cathodal-inhibitory response in healthy individuals is in line with the proposed up- and downregulation of neural excitability frequently observed in the motor system.^50^ However, polarity specific modulation of behavioral performance has been less consistent for cognition. This is likely explained by higher redundancy within the neural networks supporting cognitive functions, rendering them less malleable to effects of inhibitory tDCS.^51^ Hence, our results contribute to the ongoing debate if tDCS can modulate cognition in a polarity specific way and strengthens the assumption that bilateral SPL is critically involved in IPS in healthy individuals.

In pwRMS, Bayesian modeling provided strong evidence for a double dissociation of tDCS effects relative to healthy controls, which emphasizes that stimulation effects in pwRMS cannot easily be derived from outcomes in neurotypical individuals. Most strikingly, however, patients with more pronounced impairment responded more favorably to cathodal tDCS, while less impaired patients showed a beneficial response to anodal tDCS. Overall, this polarity by impairment interaction in pwRMS is in line with imaging studies, suggesting a gradient of neural compensation and dysfunction that depends on the level of impairment in specific tasks.^28–33^ Notably, similar mechanisms have been suggested for normal aging, where increased task-related activity in fronto-parietal regions can effectively compensate for structural and functional brain changes to a certain degree of task difficulty, while this may not be possible with more pronounced neurodegeneration.^52,53^ Hence, facilitation of compensatory processes by anodal tDCS in earlier stages of the disease, and inhibition of dysfunctional processes later (e.g., increased neural noise due to reduced lateral inhibition or disconnection) are likely candidates to explain our findings in pwRMS. Importantly, our sample was comprised of mild-to-moderately impaired patients, suggesting that the transition between the two processes may happen relatively early. This is also in line with one of the few longitudinal fMRI studies in pwRMS that demonstrated an association of increased parietal activity over time with SDMT performance declines.^54^ However, in the future, this hypothesis could be investigated more directly by administering bilateral focal SPL-tDCS concurrently with fMRI adaptation of the SDMT, to confirm the proposed mechanisms.^12,55^

There are some limitations to this trial. First, our sample was relatively small and for feasibility reasons, we only recruited patients with mild-to-moderate levels of functional impairment. Nonetheless, even in this sample, we observed a statistically sound interaction between group, polarity and stimulation condition and a dissociation between effects of excitatory and inhibitory tDCS that was explained by baseline SDMT impairment. While this suggests that patients with higher levels of impairment and brain dysfunction may benefit from cathodal SPL-tDCS, this needs to be confirmed in the future. Moreover, the double dissociation of tDCS effects between healthy individuals and pwRMS likely reflects study specific distributions of (mild-to-moderate) baseline impairment in the patients and may not generalize to samples includingh more impaired patients. Nonetheless, the impairment profile of patients recruited for this study highlights that the transition from compensatory to dysfunctional activity in the SPL may happen relatively early in the course of the disease and is therefore suited to inform future tDCS treatment decisions.

Second, because our proof-of-principle study did not involve concurrent fMRI, the proposed mechanisms by which anodal or cathodal tDCS improved SDMT performance remain to be determined. Importantly, the experimental SDMT used in this study was designed to require minimal adaptations for future tDCS-fMRI implementations, to investigate the neural mechanisms of impairment (during sham tDCS) and neuromodulation by active tDCS (anodal vs. cathodal).

Third, based on the limited number of studies that investigated tDCS effects on cognition in pwRMS and to prevent inducing maladaptive neuroplastic adaptation, we opted for a cross-over design with a single active tDCS session. Because effects of single tDCS sessions are transient and reversible, this assured that no long-term negative effects would be induced.^50^ Notably, more pronounced and longer lasting tDCS effects have been confirmed for learning compared to cognitive tasks and for multisession tDCS in combination with behavioral training or therapy.^25,27,56–58^ In this context, it is acknowledged that our study only provides proof-of-principle evidence and we abstain from claiming that the observed effects are clinically relevant. Nonetheless, the direction of effects is plausible regarding imaging findings in pwRMS and theoretical accounts of tDCS effects. Thus, we provide a rationale and guidance for future intervention studies that combine traditional neurocognitive or gaming-based interventions aimed at improving IPS with tDCS, which may be suited to induce more pronounced and lasting effects on IPS than those observed in the present study.^59,60^

Overall, we provide strong evidence for causal involvement of SPL in IPS in health and disease. The double dissociation of tDCS effects in pwRMS and healthy controls and variability of stimulation effects within the patient groups highlights the need to tailor stimulation protocols based in relevant participant-dependent factors. Our own data suggest that performance on the clinical version of the SDMT may be suited to inform tDCS treatment decisions in pwRMS and that the degree of IPS impairment reflects compensatory or dysfunctional neuroplastic processes in the SPL, that can be modulated by tDCS in a polarity specific way and that the transition between compensation and dysfunctional breakdown of neural networks supporting cognition may happen early in the course of disease. Our findings will help to inform future clinical trials aimed at improving impaired IPS in a more sustained way by combining repeated tDCS sessions and behavioral interventions.

## MATERIALS AND METHODS

This study was designed as a registered (NCT04667221), prospective, randomized, three-way blinded (i.e., participants, staff conducting the experiment and evaluating outcomes), sham-tDCS controlled, cross-over trial with a mixed-factors design and eight arms, representing the between-subjects factors group (pwRMS, healthy controls; N=32/group; **Table 1**) and tDCS polarity (anodal, cathodal), and the within-subjects factor stimulation (active, sham tDCS). All participants completed baseline assessments, including comprehensive neuropsychological assessments, followed by the experimental cross-over phase during which the computerized SDMT was completed twice. CONSORT table is provided in **Table S14** and CONSORT flow charts are in **Figure S2** for pwRMS and **Figure S3** for healthy controls.

### Participants

Thirty-two pwRMS (male/female: 11/21; Mean±SD age: 46.9±11.3 years; ethnicity was not assessed) were recruited via the outpatient clinic of the University Medicine Greifswald and selfhelp groups. Inclusion criteria comprised age≥18 years, a specialist confirmed diagnosis of RMS based on the revised McDonald criteria, no acute relapse and/or application of corticosteroids within the last three months prior to the experimental intervention, normal or corrected to normal vision, sufficient hand motor function to respond by button press on a computer keyboard, and being a native German speaker.^61^ Pre-established exclusion criteria included other major medical or neuropsychiatric diseases (e.g., major depression) and contraindications for tDCS (e.g., metal implants, prior medical procedures involving head or spinal cord, head trauma with unconsciousness, history of epilepsy, convulsions, seizures, migraine, current pregnancy).^13^ The sample size was based on previous cross-over tDCS trials from our group in neurological populations (Note: these trials only involved comparison of anodal and sham tDCS, and larger effects were expected for the between group comparison of anodal and cathodal tDCS).^62,63^

Thirty-two healthy individuals were recruited from the local community and retrospectively matched to individual patients on a 1:1 basis by age and sex (male/female: 11/21; Mean±SD age: 46.3±11.0).

All participants provided written informed consent prior to study inclusion, completed a demographic questionnaire, a comprehensive neuropsychological baseline examination and were screened for depression and anxiety. Characteristics of patients and healthy controls, results of neuropsychological baseline assessments and additional questionnaires are detailed in **Table 1**.

The study was approved by the Medical Ethics Committee of the University Medicine Greifswald, conducted in accordance with the Helsinki Declaration and registered with ClinTrials.gov (NCT04667221).

After completing baseline assessments, patients were randomly assigned to the stimulation arms by a computer-generated list (i.e., 32 codes assigning participants to specific experimental protocols, including tDCS polarity, order and right/left coding of response buttons for the experimental task) and participated in the experimental cross-over phase of the study. The order of sham and active stimulation was counterbalanced across the groups, i.e., half of the participants received either active or sham tDCS during the first session (**Figure 1A**). Healthy controls received the same stimulation protocol as patients they were matched to. Two pwRMS in the anodal tDCS arm had to be excluded due to technical errors (i.e., software issues that prevented starting the experimental paradigm) during one of their stimulation sessions.

### Experimental SDMT

To minimize reliance on fine motor skills required by the original paper-and-pencil SDMT, we developed a computerized version that resembled previous MRI adaptations with a forced-choice button press response mode.^34,48^ This also allowed creating a longer version of the task, to enhance statistical power to detect potential stimulation effects. The experimental SDMT paradigm was presented using NBS Presentation® (Version 19.0, Neurobehavioral Systems, Inc., Berkeley, CA) During the experimental sessions, participants completed nine experimental blocks, each with a fixed duration of 90 sec (**Figure 1B**). Each block comprised nine unique and visually distinct symbols and digits (1-9) and a legend showing nine symbol-digit combinations (**Figures 2**). During each trial, the legend was shown at the top of the screen; at the bottom of the screen, a single symbol-digit-combination was displayed (probe), which was either coherent or incoherent with the legend. Participants were asked to indicate coherence of the probe with the legend by button-presses with left and right index finger (“M” and “Y” keys) on a QWERTZ keyboard. Incoherent probes within blocks were created by systematically shifting the symbol-digit-combinations. For each coherent combination, eight unique shifted “incoherent” combinations were created (i.e., symbol and digit+1-8). During each task block, one shifted set was randomly chosen, and coherent and incoherent symbol-digit-combinations were pseudo-randomly presented, so that the same digit or symbol was not presented twice in a row. After all pairings were presented, another shifted set was randomly chosen. This assured that approximately the same number of coherent and incoherent trials were presented. The task was self-paced, and the next trial appeared immediately after each response. Participants were instructed to respond as quickly as possible, without making mistakes. No immediate feedback on performance was provided during or after each block. The maximum number of trials per block was 144, which was unattainable within the 90 sec block duration. Short breaks were interspersed in between blocks (max. 60 sec) and participants could proceed with the next block at their own pace during this time.

### Transcranial direct current stimulation

Stimulation was administered with a Neuroelectrics Starstim8 direct current stimulator using Pistim electrodes (radius=1cm) and a 3×1 set-up (i.e., a central active electrode, and three surrounding reference electrodes). Circular inserts in an EEG-cap secured the electrodes on the head. Center electrodes were positioned bilaterally over the SPL based on 10-10 EEG coordinates (i.e., P1/P2). Return electrodes were positioned in a circular way around the anodes (CP2, P5, PO4, CP1, P6, PO3). This set-up was chosen during the planning stage of the project based on current flow simulations using standard SimNIBS parameters and a MNI152 standard brain, that demonstrated peak current intensities in the superior parietal cortex (BA7) and additional current flow to surrounding and deeper brain regions, including the precuneus (**Figure 3**).^64,65^ Stimulation commenced briefly prior to the start of the experimental sessions and was applied with 1.5 mA for 20 minutes with a 20s ramp-up and -down at the beginning and the end (active tDCS), or was ramped-up and -down over 40s (sham tDCS). This procedure which has been shown to ensure participant blinding for focal set-ups.^37,39,41^ Staff administering the stimulation were blinded using predefined stimulation codes that concealed the applied stimulation type.

An adverse effects questionnaire was completed by the participants after each stimulation session^13^ Potential unspecific effects of tDCS were assessed with the Positive and Negative Affect Schedule (PANAS) before and after each session.^44^ After the second experimental session, subjects were asked if they believed that active stimulation was administered during the first or the second tDCS session to assess blinding integrity. Potential responses included “I do not know”, “During the first session”, or “During the second session”.

### Outcome measures

Response latency for correct responses was our predefined primary outcome measure (NCT04667221). We also inspected potential effects of tDCS on accuracy as secondary outcome. Response latency data were preprocessed in three steps: (1) All trials with incorrect responses were excluded. (2) Only trials with response latencies between 0.2 and 6s were included, in line with the upper limit that was used by previous studies that used similar designs.^42,66,67^ (3) Trials with response latencies within the interval “median±3×median absolute deviation”, computed on a subject and session basis, were included in the analysis. This approach is more robust than intervals based on means and standard deviations.^68^ Across sessions and stimulation groups, 5.65-7.06% of trials were excluded (**Table S15**).

### Statistical Analyses

Statistical analyses were conducted in a Bayesian framework using R and the *brms* package.^69,70^ Data were analyzed using generalized linear mixed models (GLMMs) using a logit-link function for response accuracy, and a lognormal-link function for response latency analyses for binomial and skewed data distributions, respectively. The response latency model was truncated to match the range of possible response time outcomes fixed by our filtering procedure. Three models were fitted for each analyses to find the best fitting model for our data: (1) An intercept only model; (2) a covariates model to correct for potential session effects, motor slowing via the TMT-A and baseline performance on the paper-pencil SDMT;^42^ (3) A full model comprising the covariates as specified above, and variables of interest (i.e., stimulation polarity [anodal, cathodal], group [pwRMS, controls], and stimulation type [active, sham]), as well as their interactions. All models included a group-level subject intercept (random intercept) to correct for individual performance levels. To establish the models that best explained our data, models were compared using the widely applicable information criterion (WAIC), which is a fully Bayesian extension of the Akaike information criterion (AIC), and the estimated log pointwise predictive density of the WAIC (ELPD WAIC). Difference between ELPD WAIC scores larger than 1.96 times than the standard error of the standard error of the ELPD difference have been suggested to be meaningful.^43^ For each model, 3000 samples per chain with eight chains were drawn after warm-up, resulting in overall 24,000 draws using the Hamiltonian Monte Carlo Algorithm.^71^ Due to convergence issues, low effective sampling size and high rhat-values, reaction time models had to be run with 5000 samples after warm-up with eight chains, i.e., 40,000 draws. Furthermore, the random intercept of the intercept model required a soft sum to zero constraint to converge.^72^

An additional analysis investigated if the degree of IPS impairment in pwRMS (quantified by the clinical SDMT administered during the baseline assessments) was associated with stimulation response (i.e., reduced or increased response latency in the respective tDCS conditions). For that, the full reaction time model was extended: Baseline performance on the paper pencil SDMT, group, stimulation polarity and stimulation type, as well as their interactions were added as population level effects. Additionally, we corrected for TMT-A performance and the session effect.^42^ Subjects were added as group-level intercepts. To ensure the validity of this analysis and enhance clinical relevance of potential outcomes, we considered two different predictors: (1) study specific raw scores of the baseline SDMT in our sample, (2) age- and education corrected norms of the SDMT.^34^

To analyze self-reported adverse effects and PANAS data, we built two GLMMs for each outcome. (1) A simple model that assumed that the effect of active stimulation compared to sham is the same in all experimental groups. (2) A more complex model that allowed the effect of active stimulation to be distinct, which allows modeling population specific vulnerability to adverse effects.

For the PANAS data, negative and positive items were summed to build a sum-score for each dimension (i.e., positive or negative valence). We fitted a hurdle Gaussian model, since negative affect included many sum-scores of 0, but responses were otherwise normally distributed.^73^ The full model included factors for stimulation polarity, group, stimulation type, time point (pre-, post tDCS), and valence (positive, negative), as well as all interactions. The simple model, however, only included interactions between stimulation type, subject group, time point and valence, and the interaction between active stimulation and time point. Additionally, we modeled the hurdle parameter separately for positive and negative affect, since zeros were only found in the measures of negative affect. The PANAS models included a group level intercept for subjects.

Adverse effects data were modeled with a cumulative model assuming a continuous latent variable (strength of adverse effect) that was measured using categorical responses (“none”, “mild”, “moderate”, “strong”).^74^ The full model included stimulation polarity, stimulation type, and group, as well as all their interactions. The simple model only considered interactions between stimulation polarity and group. Both models included a population-level factor for sensation types (i.e., itching, pain, burning, heat, metallic taste, fatigue, other) and group-level intercepts for sensation types and subjects.

To investigate blinding of participants, we used categorical models to interrogate the participants’ responses to the question: “During which session do you think you were stimulated?” With categorical models, intercepts represent the shift from one category to another, e.g., the transition from the answer “do not know” to “I think, I was stimulated during the first session”. Factors in the models shift the location of these transitions, so that more participants fall into a certain category. Again, we fitted multiple models to describe the data. A full model that included the session when stimulation was applied the group and the stimulation polarity, as well as their interactions. A simple model that only included the interaction of group and the stimulation polarity, and a simple effect for the active stimulation session. Thus, we were able to model if the correct response is associated with the participants’ response and consistency of this relationship across groups. Model comparisons for the adverse effects, PANAS, and blinding models were investigated with WAIC and ELPD WAIC as criteria for model comparisons.

### Prior choices

In a Bayesian framework, priors must be chosen to represent expected values the model parameters may take before the current data was considered. Experimental data shifts these priors to represent the expectations after considering data. Priors were chosen to be weakly informative, i.e., population-level parameters were normally distributed with a mean of 0 and standard deviation of 1, and group-level standard deviations were set to an exponential distribution with rate of 2 in all models. Note that the blinding assessment did not include group level intercept, since only one answer per participant was collected.

### Hypothesis testing

Bayesian hypotheses testing used evidence ratios. For directed hypothesis, an evidence ratio is the number of draws in the direction of an effect relative to the number of draws in the opposite direction, i.e., the evidence ratio for a learning effect across sessions would be computed: 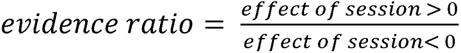. Thus, evidence ratios indicate the amount of evidence in favor of a hypothesis compared to the direct counter hypothesis. Evidence ratios were computed for the model parameters and post-hoc comparisons. The theoretical range of evidence ratios is 0 to ∞, whereby evidence ratios of ∞ indicate that all posterior draws were in favor of the tested hypotheses. For point-hypotheses, i.e., tests for equality, evidence ratios indicate an inor decrease of evidence compared to the prior model.^70^ Simulations with linear and logistic regression models have found that evidence ratios of 19 and 39 correspond to one- and two-sided hypothesis tests with an alpha-level of 0.05.^75^ Note that we are unaware of formal comparisons between evidence ratios and p-values in generalized linear mixed models. Post-hoc comparisons were always comprised of within subjects comparisons of active session to the respective sham sessions. For accuracy and response latency models, differences between the sessions were tested, while adverse effects, blinding, and PANAS models tested equality of sessions.

## Data availability

The experimental paradigm and data, as well as scripts for data analyses are available at open science framework (OSF).

## REFERENCES

1. Jakimovski D, et al. Multiple sclerosis. The Lancet. 2024;403(10422):183–202. 10.1016/S0140-6736(23)01473-3

2. Brownlee WJ, Hardy TA, Fazekas F, Miller DH. Diagnosis of multiple sclerosis: progress and challenges. The Lancet. 2017;389(10076):1336–1346. 10.1016/S0140-6736(16)30959-X

3. Benedict RHB, Amato MP, DeLuca J, Geurts JJG. Cognitive impairment in multiple sclerosis: clinical management, MRI, and therapeutic avenues. The Lancet Neurology. 2020;19(10):860–871. 10.1016/S1474-4422(20)30277-5

4. Kobelt G et al. New insights into the burden and costs of multiple sclerosis in Europe. Multiple Sclerosis (Houndmills, Basingstoke, England). 2017;23(8):1123–1136. 10.1177/1352458517694432

5. Deloire M et al. Early cognitive impairment in multiple sclerosis predicts disability outcome several years later. Multiple Sclerosis Journal. 2010;16(5):581–587. 10.1177/1352458510362819

6. Patti F et al. Prevalence and incidence of cognitive impairment in multiple sclerosis: a population-based survey in Catania, Sicily. Journal of Neurology. 2015;262(4):923–930. 10.1007/s00415-015-7661-3

7. Ruet A et al. Cognitive impairment, health-related quality of life and vocational status at early stages of multiple sclerosis: a 7-year longitudinal study. Journal of Neurology. 2013;260(3):776–784. 10.1007/s00415-012-6705-1

8. Bergmann C et al. Multiple sclerosis and quality of life: The role of cognitive impairment on quality of life in people with multiple sclerosis. Multiple Sclerosis and Related Disorders. 2023;79:104966. 10.1016/j.msard.2023.104966

9. Sumowski JF et al. Cognition in multiple sclerosis. Neurology. 2018;90(6):278–288. 10.1212/WNL.0000000000004977

10. Costa SL, Genova HM, DeLuca J, Chiaravalloti ND. Information processing speed in multiple sclerosis: Past, present, and future. Multiple Sclerosis Journal. 2017;23(6):772–789. 10.1177/1352458516645869

11. Hauser SL, Cree BAC. Treatment of Multiple Sclerosis: A Review. The American Journal of Medicine. 2020;133(12):1380–1390.e2. 10.1016/j.amjmed.2020.05.049

12. Meinzer M et al. Investigating the neural mechanisms of transcranial direct current stimulation effects on human cognition: current issues and potential solutions. Frontiers in Neuroscience. 2024 [accessed 2024 Jun 25];18. https://www.frontiersin.org/journals/neuroscience/articles/10.3389/fnins.2024.1389651/full. 10.3389/fnins.2024.1389651

13. Antal A et al. Low intensity transcranial electric stimulation: Safety, ethical, legal regulatory and application guidelines. Clinical Neurophysiology: Official Journal of the International Federation of Clinical Neurophysiology. 2017;128(9):1774–1809. 10.1016/j.clinph.2017.06.001

14. Narmashiri A, Akbari F. The Effects of Transcranial Direct Current Stimulation (tDCS) on the Cognitive Functions: A Systematic Review and Meta-analysis. Neuropsychology Review. 2023 Dec 7 [accessed 2023 Dec 8]. 10.1007/s11065-023-09627-x. 10.1007/s11065-023-09627-x

15. Perceval G, Flöel A, Meinzer M. Can transcranial direct current stimulation counteract age-associated functional impairment? Neuroscience & Biobehavioral Reviews. 2016;65:157–172. 10.1016/j.neubiorev.2016.03.028

16. Cammisuli DM et al. Transcranial Direct Current Stimulation (tDCS) as a Useful Rehabilitation Strategy to Improve Cognition in Patients With Alzheimer’s Disease and Parkinson’s Disease: An Updated Systematic Review of Randomized Controlled Trials. Frontiers in Neurology. 2022 [accessed 2024 Jul 26];12. https://www.frontiersin.org/journals/neurology/articles/10.3389/fneur.2021.798191/full. 10.3389/fneur.2021.798191

17. Chen J et al. Transcranial Direct Current Stimulation Enhances Cognitive Function in Patients with Mild Cognitive Impairment and Early/Mid Alzheimer’s Disease: A Systematic Review and Meta-Analysis. Brain Sciences. 2022;12(5):562. 10.3390/brainsci12050562

18. Ciullo V et al. Transcranial Direct Current Stimulation and Cognition in Neuropsychiatric Disorders: Systematic Review of the Evidence and Future Directions. The Neuroscientist. 2021;27(3):285–309. 10.1177/1073858420936167

19. Flöel A. tDCS-enhanced motor and cognitive function in neurological diseases. NeuroImage. 2014;85:934–947. (Neuro-enhancement). 10.1016/j.neuroimage.2013.05.098

20. Hiew S, Nguemeni C, Zeller D. Efficacy of transcranial direct current stimulation in people with multiple sclerosis: a review. European Journal of Neurology. 2022;29(2):648–664. 10.1111/ene.15163

21. Hsu W-Y et al. Effects of Transcranial Direct Current Stimulation on Cognition, Mood, Pain, and Fatigue in Multiple Sclerosis: A Systematic Review and Meta-Analysis. Frontiers in Neurology. 2021 [accessed 2024 Jul 22];12. https://www.frontiersin.org/journals/neurology/articles/10.3389/fneur.2021.626113/full. 10.3389/fneur.2021.626113

22. Grigorescu C et al. Effects of Transcranial Direct Current Stimulation on Information Processing Speed, Working Memory, Attention, and Social Cognition in Multiple Sclerosis. Frontiers in Neurology. 2020 [accessed 2024 Jun 28];11. https://www.frontiersin.org/journals/neurology/articles/10.3389/fneur.2020.545377/full. 10.3389/fneur.2020.545377

23. Mattioli F et al. Neuroenhancement through cognitive training and anodal tDCS in multiple sclerosis. Multiple Sclerosis Journal. 2016;22(2):222–230. 10.1177/1352458515587597

24. Allman C et al. Ipsilesional anodal tDCS enhances the functional benefits of rehabilitation in patients after stroke. Science Translational Medicine. 2016;8(330):330re1-330re1. 10.1126/scitranslmed.aad5651

25. Perceval G et al. Multisession transcranial direct current stimulation facilitates verbal learning and memory consolidation in young and older adults. Brain and Language. 2020;205:104788. 10.1016/j.bandl.2020.104788

26. Yu T-H, Wu Y-J, Chien M-E, Hsu K-S. Multisession Anodal Transcranial Direct Current Stimulation Enhances Adult Hippocampal Neurogenesis and Context Discrimination in Mice. The Journal of Neuroscience: The Official Journal of the Society for Neuroscience. 2023;43(4):635–646. 10.1523/JNEUROSCI.1476-22.2022

27. Meinzer M, Darkow R, Lindenberg R, Flöel A. Electrical stimulation of the motor cortex enhances treatment outcome in post-stroke aphasia. Brain: A Journal of Neurology. 2016;139(Pt 4):1152–1163. 10.1093/brain/aww002

28. Rocca MA et al. Task- and resting-state fMRI studies in multiple sclerosis: From regions to systems and time-varying analysis. Current status and future perspective. NeuroImage: Clinical. 2022;35:103076. 10.1016/j.nicl.2022.103076

29. Rocca MA et al. Adaptive functional changes in the cerebral cortex of patients with nondisabling multiple sclerosis correlate with the extent of brain structural damage. Annals of Neurology. 2002;51(3):330–339. 10.1002/ana.10120

30. Rocca MA et al. Abnormal connectivity of the sensorimotor network in patients with MS: A multicenter fMRI study. Human Brain Mapping. 2009;30(8):2412–2425. 10.1002/hbm.20679

31. Wegner C et al. Relating functional changes during hand movement to clinical parameters in patients with multiple sclerosis in a multi-centre fMRI study. European Journal of Neurology. 2008;15(2):113–122. 10.1111/j.1468-1331.2007.02027.x

32. Rocca MA et al. Pyramidal tract lesions and movement-associated cortical recruitment in patients with MS. NeuroImage. 2004;23(1):141–147. 10.1016/j.neuroimage.2004.05.005

33. Lenzi D et al. Effect of corpus callosum damage on ipsilateral motor activation in patients with multiple sclerosis: A functional and anatomical study. Human Brain Mapping. 2007;28(7):636–644. 10.1002/hbm.20305

34. Smith A. Symbol digit modalities test (SDMT) manual (revised) Western psychological services. Los Angeles. 1982:10.

35. Grothe M et al. Functional representation of the symbol digit modalities test in relapsing remitting multiple sclerosis. Multiple Sclerosis and Related Disorders. 2020;43:102159. 10.1016/j.msard.2020.102159

36. Grothe M et al. Performance in information processing speed is associated with parietal white matter tract integrity in multiple sclerosis. Frontiers in Neurology. 2022 [accessed 2024 Jul 22];13. https://www.frontiersin.org/journals/neurology/articles/10.3389/fneur.2022.982964/full. 10.3389/fneur.2022.982964

37. Gbadeyan O, Steinhauser M, McMahon K, Meinzer M. Safety, Tolerability, Blinding Efficacy and Behavioural Effects of a Novel MRI-Compatible, High-Definition tDCS Set-Up. Brain Stimulation. 2016;9(4):545–552. 10.1016/j.brs.2016.03.018

38. Lischke A et al. Improving mentalizing deficits in older age with region-specific transcranial direct current stimulation. GeroScience. 2024 Jun 15 [accessed 2024 Jul 22]. 10.1007/s11357-024-01206-z. 10.1007/s11357-024-01206-z

39. Martin AK, Huang J, Hunold A, Meinzer M. Dissociable Roles Within the Social Brain for Self–Other Processing: A HD-tDCS Study. Cerebral Cortex. 2019;29(8):3642–3654. 10.1093/cercor/bhy238

40. Martin AK et al. The Right Temporoparietal Junction Is Causally Associated with Embodied Perspective-taking. Journal of Neuroscience. 2020;40(15):3089–3095. 10.1523/JNEUROSCI.2637-19.2020

41. Riemann S et al. The role of frontal cortex in novel-word learning and consolidation: Evidence from focal transcranial direct current stimulation. Cortex. 2024;177:15–27. 10.1016/j.cortex.2024.05.004

42. Genova HM et al. Examination of processing speed deficits in multiple sclerosis using functional magnetic resonance imaging. Journal of the International Neuropsychological Society. 2009;15(3):383–393. 10.1017/S1355617709090535

43. Vehtari A, Gelman A, Gabry J. Practical Bayesian model evaluation using leave-one-out cross-validation and WAIC. Statistics and Computing. 2017;27(5):1413–1432. 10.1007/s11222-016-9696-4

44. Watson D, Clark LA, Tellegen A. Development and validation of brief measures of positive and negative affect: The PANAS scales. Journal of Personality and Social Psychology. 1988;54(6):1063–1070. 10.1037/0022-3514.54.6.1063

45. Breakspear M. Dynamic models of large-scale brain activity. Nature neuroscience. 2017;20(3):340–352.

46. Kang J et al. Effects and safety of transcranial direct current stimulation on multiple health outcomes: an umbrella review of randomized clinical trials. Molecular Psychiatry. 2024 May 30:1–13. 10.1038/s41380-024-02624-3

47. Fertonani A, Miniussi C. Transcranial Electrical Stimulation: What We Know and Do Not Know About Mechanisms. The Neuroscientist. 2017;23(2):109–123. 10.1177/1073858416631966

48. Silva PHR, Spedo CT, Barreira AA, Leoni RF. Symbol Digit Modalities Test adaptation for Magnetic Resonance Imaging environment: A systematic review and meta-analysis. Multiple Sclerosis and Related Disorders. 2018;20:136–143. 10.1016/j.msard.2018.01.014

49. Koenigs M, Barbey AK, Postle BR, Grafman J. Superior Parietal Cortex Is Critical for the Manipulation of Information in Working Memory. Journal of Neuroscience. 2009;29(47):14980–14986. 10.1523/JNEUROSCI.3706-09.2009

50. Stagg CJ, Antal A, Nitsche MA. Physiology of Transcranial Direct Current Stimulation. The Journal of ECT. 2018;34(3):144. 10.1097/YCT.0000000000000510

51. Jacobson L, Koslowsky M, Lavidor M. tDCS polarity effects in motor and cognitive domains: a meta-analytical review. Experimental Brain Research. 2012;216(1):1–10. 10.1007/s00221-011-2891-9

52. Oosterhuis EJ, Slade K, May PJC, Nuttall HE. Toward an Understanding of Healthy Cognitive Aging: The Importance of Lifestyle in Cognitive Reserve and the Scaffolding Theory of Aging and Cognition. The Journals of Gerontology: Series B. 2023;78(5):777–788. 10.1093/geronb/gbac197

53. Reuter-Lorenz PA, Park DC. How does it STAC up? Revisiting the scaffolding theory of aging and cognition. Neuropsychology Review. 2014;24(3):355–370. 10.1007/s11065-014-9270-9

54. Loitfelder M et al. Brain Activity Changes in Cognitive Networks in Relapsing-Remitting Multiple Sclerosis – Insights from a Longitudinal fMRI Study. PLOS ONE. 2014;9(4):e93715. 10.1371/journal.pone.0093715

55. Niemann F et al. Neuronavigated Focalized Transcranial Direct Current Stimulation Administered During Functional Magnetic Resonance Imaging. Journal of Visual Experiments. in press. DOI: 10.3791/67155

56. Simonsmeier BA et al. Electrical brain stimulation (tES) improves learning more than performance: A meta-analysis. Neuroscience & Biobehavioral Reviews. 2018;84:171–181. 10.1016/j.neubiorev.2017.11.001

57. Song S et al. Effects of single-session versus multi-session non-invasive brain stimulation on craving and consumption in individuals with drug addiction, eating disorders or obesity: A metaanalysis. Brain Stimulation. 2019;12(3):606–618. 10.1016/j.brs.2018.12.975

58. Antonenko D et al. Microstructural and functional plasticity following repeated brain stimulation during cognitive training in older adults. Nature Communications. 2023;14:3184. 10.1038/s41467-023-38910-x

59. Chiaravalloti ND et al. The efficacy of speed of processing training for improving processing speed in individuals with multiple sclerosis: a randomized clinical trial. Journal of Neurology. 2022;269(7):3614–3624. 10.1007/s00415-022-10980-9

60. Dye MWG, Green CS, Bavelier D. Increasing Speed of Processing With Action Video Games. Current Directions in Psychological Science. 2009;18(6):321–326. 10.1111/j.1467-8721.2009.01660.x

61. Thompson AJ et al. Diagnosis of multiple sclerosis: 2017 revisions of the McDonald criteria. The Lancet Neurology. 2018;17(2):162–173. 10.1016/S1474-4422(17)30470-2

62. Meinzer M et al. Transcranial direct current stimulation in mild cognitive impairment: Behavioral effects and neural mechanisms. Alzheimer’s & Dementia. 2015;11(9):1032–1040. 10.1016/j.jalz.2014.07.159

63. Darkow R et al. Transcranial direct current stimulation effects on neural processing in poststroke aphasia. Human Brain Mapping. 2017;38(3):1518–1531. 10.1002/hbm.23469

64. Saturnino GB et al. SimNIBS 2.1: A Comprehensive Pipeline for Individualized Electric Field Modelling for Transcranial Brain Stimulation. In: Makarov S, Horner M, Noetscher G, editors. Brain and Human Body Modeling: Computational Human Modeling at EMBC 2018. Springer; 2019. http://www.ncbi.nlm.nih.gov/books/NBK549569/

65. Thielscher A, Antunes A, Saturnino GB. Field modeling for transcranial magnetic stimulation: A useful tool to understand the physiological effects of TMS? In: 2015 37th Annual International Conference of the IEEE Engineering in Medicine and Biology Society (EMBC). 2015. p 222–225. 10.1109/EMBC.2015.7318340

66. Gawryluk JR, Mazerolle EL, Beyea SD, D’Arcy RCN. Functional MRI activation in white matter during the Symbol Digit Modalities Test. Frontiers in Human Neuroscience. 2014 [accessed 2024 May 13];8. https://www.frontiersin.org/articles/10.3389/fnhum.2014.00589. 10.3389/fnhum.2014.00589

67. Leavitt VM et al. Altered effective connectivity during performance of an information processing speed task in multiple sclerosis. Multiple Sclerosis Journal. 2012;18(4):409–417. 10.1177/1352458511423651

68. Rousseeuw PJ, Hubert M. Robust statistics for outlier detection. WIREs Data Mining and Knowledge Discovery. 2011;1(1):73–79. 10.1002/widm.2

69. R Core Team. R: A Language and Environment for Statistical Computing. R Foundation for Statistical Computing; 2022. https://www.R-project.org/

70. Bürkner P-C. brms: An R Package for Bayesian Multilevel Models Using Stan. Journal of Statistical Software. 2017;80(1):1–28. 10.18637/jss.v080.i01

71. Hoffman MD, Gelman A. The No-U-Turn Sampler: Adaptively Setting Path Lengths in Hamiltonian Monte Carlo. Journal of Machine Learning Research. 2014;15(47):1593–1623.

72. STAN Forum. Test: Soft vs Hard sum-to-zero constrain + choosing the right prior for soft constrain - Modeling. The Stan Forums. 2018 May 13 [accessed 2024 Aug 8]. https://discourse.mc-stan.org/t/test-soft-vs-hard-sum-to-zero-constrain-choosing-the-right-prior-for-soft-constrain/3884?page=2

73. Welsh AH, Cunningham RB, Donnelly CF, Lindenmayer DB. Modelling the abundance of rare species: statistical models for counts with extra zeros. Ecological Modelling. 1996;88(1):297–308. 10.1016/0304-3800(95)00113-1

74. Bürkner P-C, Vuorre M. Ordinal Regression Models in Psychology: A Tutorial. Advances in Methods and Practices in Psychological Science. 2019;2(1):77–101. 10.1177/2515245918823199

75. Makowski D, Ben-Shachar MS, Chen SHA, Lüdecke D. Indices of Effect Existence and Significance in the Bayesian Framework. Frontiers in Psychology. 2019 [accessed 2023 Feb 7];10. https://www.frontiersin.org/articles/10.3389/fpsyg.2019.02767

