## Supplementary material for "Information processing speed modulation by electrical brain stimulation in multiple sclerosis: Towards individually-tailored protocols": Figure S1

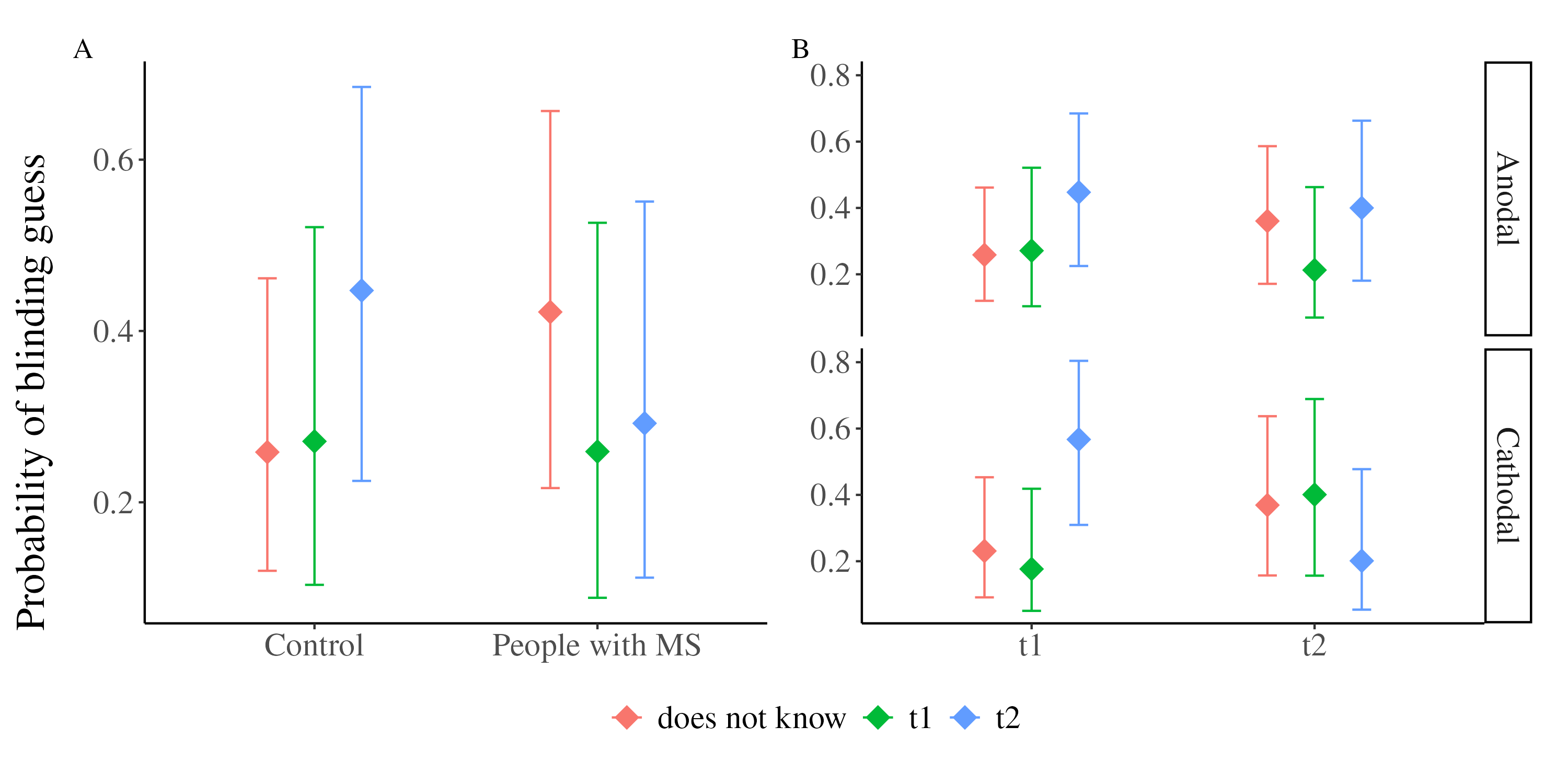

**Supplementary Figure 1. Conditional Effects of the blinding model.** Illustration of the effect (A) that patients were more likely to answer that they do not know, when they were stimulated and (B) that people that were stimulated during the second session with cathodal stimulation were more likely to answer that they do not know when they were stimulated.
