## Supplementary figures and images for "Information processing speed modulation by electrical brain stimulation in multiple sclerosis: Towards individually-tailored protocols"

### Figure S2

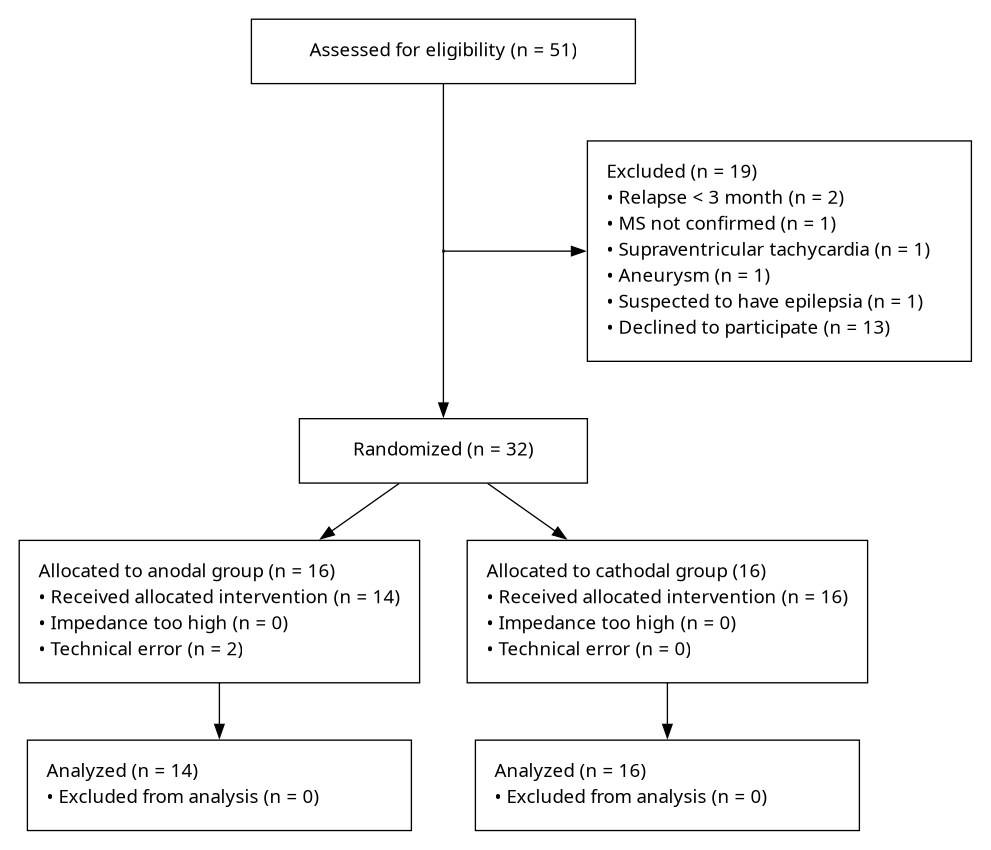

**Supplementary Figure 2. CONSORT flow-chart pwRMS.**

### Figure S3

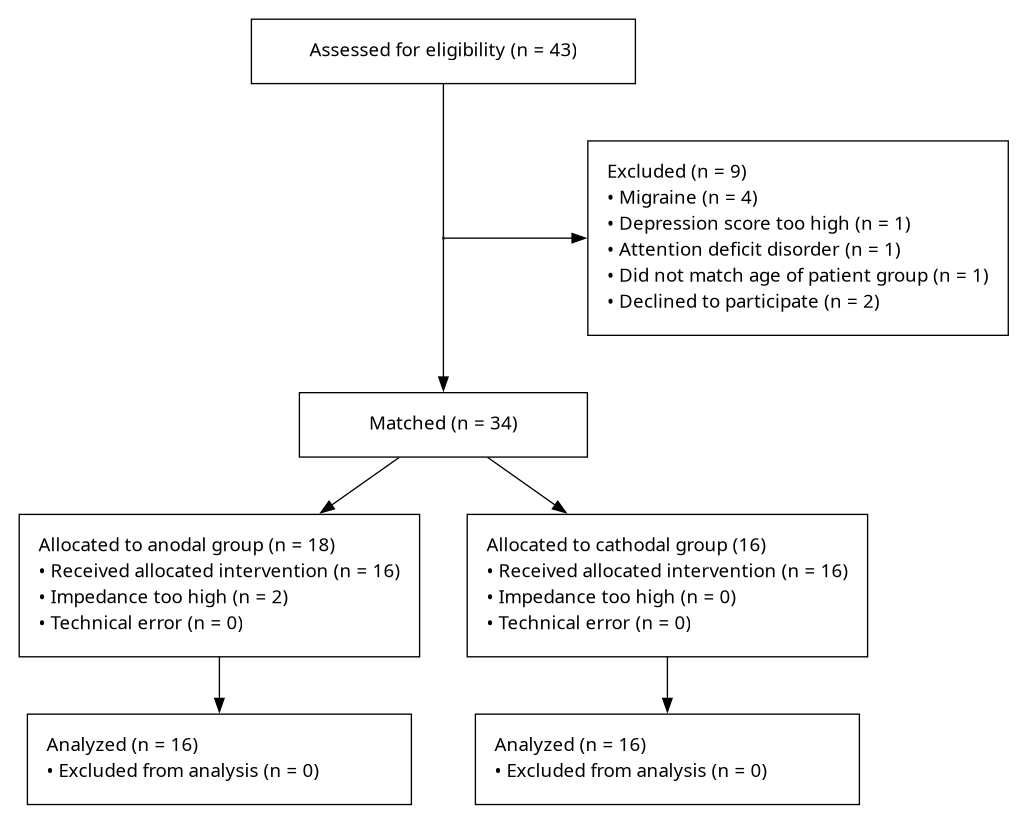
