## Supplementary material for "Information processing speed modulation by electrical brain stimulation in multiple sclerosis: Towards individually-tailored protocols": Table S1

| **Supplementary Table 2** | | | | | | | | |
| --- | --- | --- | --- | --- | --- | --- | --- | --- |
| *Summary of accuracy data* | | | | | | | | |
|  | *Control* | | | | *pwRMS* | | | |
|  | *anodal* | | *cathodal* | | *anodal* | | *cathodal* | |
|  | *M* | *SD* | *M* | *SD* | *M* | *SD* | *M* | *SD* |
| sham | 0.99 | 0.12 | 0.98 | 0.13 | 0.98 | 0.15 | 0.98 | 0.13 |
| active | 0.98 | 0.13 | 0.98 | 0.14 | 0.98 | 0.14 | 0.99 | 0.12 |
| *Note.* The table shows the mean and standard deviation of accuracy for each group, stimulation group and active condition. M=Mean, SD=Standard deviation. pwRMS=patients with relapsing multiple sclerosis | | | | | | | | |
