## Supplementary material for "Information processing speed modulation by electrical brain stimulation in multiple sclerosis: Towards individually-tailored protocols": Table S2

| **Supplementary Table 3** | | | | | | | | |
| --- | --- | --- | --- | --- | --- | --- | --- | --- |
| *WAIC comparison of response accuracy models. ELPD diff shows the difference in the ELPD WAIC-value relative to the best performing model (here: model including covariates).* | | | | | | | | |
| Model | ELPD diff | | ELPD WAIC | | pWAIC | | WAIC | |
|  | Est | *SE* | Est | *SE* | Est | *SE* | Est | *SE* |
| Covariates model^1^ | 0.00 | 0.00 | -4,866.42 | 122.35 | 48.76 | 1.62 | 9,732.85 | 244.69 |
| Full model^1, 2^ | -1.40 | 2.48 | -4,867.82 | 122.45 | 53.40 | 1.76 | 9,735.65 | 244.90 |
| Intercept only model | -8.02 | 4.14 | -4,874.44 | 122.53 | 48.99 | 1.64 | 9,748.89 | 245.07 |
| *Note.* Models are ranked according to their ELPD WAIC score, i.e., the top model reached the best quality scores, while the bottom model has the worst. While the intercept only model does not yield better model criteria (\|-8.02\| < 8.11), the simpler intercept only model is favored over the more complex covariates model or the full model. ELPD=Expected log pointwise predictive density, SE=Standard error, WAIC=Widely applicable information criterion, pWAIC=Effective number of parameters.  ^1^ Includes covariates to correct for the effects of TMT-A time, SDMT n correct at neuropsychology, and session.  ^2^ Includes effects of interest, i.e., subject group, active stimulation, and stimulation type, as well as their interactions. | | | | | | | | |
