## Supplementary material for "Information processing speed modulation by electrical brain stimulation in multiple sclerosis: Towards individually-tailored protocols": Table S4

| **Supplementary Table 5** | | | | | | | | |
| --- | --- | --- | --- | --- | --- | --- | --- | --- |
| *WAIC comparison of response latency models. ELPD diff shows the difference in the ELPD-value relative to the best performing model (here: the full model).* | | | | | | | | |
| Model | ELPD diff | | ELPD WAIC | | pWAIC | | WAIC | |
|  | Est | *SE* | Est | *SE* | Est | *SE* | Est | *SE* |
| Full model^1, 2^ | 0.00 | 0.00 | -21,282.99 | 190.79 | 70.39 | 0.48 | 42,565.98 | 381.57 |
| Covariates model^2^ | -95.72 | 13.70 | -21,378.70 | 191.43 | 66.04 | 0.45 | 42,757.41 | 382.86 |
| Intercept only model | -1,751.70 | 58.11 | -23,034.69 | 194.51 | 65.07 | 0.45 | 46,069.38 | 389.03 |
| *Note.* Models are ranked according to their ELPD WAIC score, i.e., the top model reached the best quality scores, while the bottom model has the worst. ELPD WAIC=Expected log pointwise predictive density of WAIC, ELPD diff=difference of the ELPD relative to the best performing model. SE=Standard error, WAIC=Widely applicable information criterion, pWAIC=Effective number of parameters.  ^1^ Includes effects of interest, i.e., subject group, active stimulation, and stimulation type, as well as their interactions.  ^2^ Includes covariates to correct for the effects of TMT-A time, SDMT n correct at neuropsychology, and session. | | | | | | | | |
