## Supplementary material for "Information processing speed modulation by electrical brain stimulation in multiple sclerosis: Towards individually-tailored protocols": Table S7

| **Supplementary Table 8** | | | | | | | |
| --- | --- | --- | --- | --- | --- | --- | --- |
| *Population-level effects of the exploratory response latency model with SDMT z-scores from Smith et al. (1982).* | | | | | | | |
| Parameter | Estimate | *SE* | Rhat | Bulk ESS | Tail ESS | Evidence ratio | 95% CI |
| Intercept | 0.56 | 0.04 | 1.00 | 3,884.33 | 7,416.25 | ∞^1^ | [0.47, 0.64] |
| Session | -0.12 | 0.00 | 1.00 | 51,161.18 | 25,510.37 | ∞^1^ | [-0.13, -0.12] |
| TMT-A time | 0.09 | 0.03 | 1.00 | 4,464.91 | 7,974.35 | 443.44 | [0.03, 0.15] |
| SDMT z-score (Smith et al., 1982) | -0.06 | 0.06 | 1.00 | 3,823.58 | 7,954.16 | 4.51 | [-0.18, 0.06] |
| Group | 0.09 | 0.08 | 1.00 | 3,547.80 | 7,038.11 | 6.80 | [-0.07, 0.26] |
| Stimulation type | -0.03 | 0.00 | 1.00 | 22,592.72 | 26,760.26 | ∞^1^ | [-0.04, -0.02] |
| Stimulation polarity | -0.12 | 0.06 | 1.00 | 3,004.09 | 6,136.42 | 32.00 | [-0.24, 0.01] |
| SDMT z-score (Smith et al., 1982) × Group | 0.01 | 0.08 | 1.00 | 3,639.20 | 7,722.49 | 1.22 | [-0.14, 0.16] |
| SDMT z-score (Smith et al., 1982) × Stimulation type | -0.03 | 0.01 | 1.00 | 14,099.24 | 21,756.18 | ∞^1^ | [-0.04, -0.02] |
| Group × Stimulation type | 0.02 | 0.01 | 1.00 | 15,344.33 | 22,139.87 | 50.81 | [0.00, 0.03] |
| SDMT z-score (Smith et al., 1982) × Stimulation polarity | -0.05 | 0.07 | 1.00 | 3,911.77 | 8,200.19 | 3.17 | [-0.19, 0.09] |
| Group × Stimulation polarity | -0.04 | 0.11 | 1.00 | 3,379.99 | 6,013.64 | 1.84 | [-0.26, 0.18] |
| Stimulation type × Stimulation polarity | 0.07 | 0.01 | 1.00 | 22,533.99 | 25,630.98 | ∞^1^ | [0.06, 0.08] |
| SDMT z-score (Smith et al., 1982) × Group × Stimulation type | 0.01 | 0.01 | 1.00 | 11,663.75 | 18,732.62 | 8.48 | [-0.01, 0.02] |
| SDMT z-score (Smith et al., 1982) × Group × Stimulation polarity | -0.06 | 0.09 | 1.00 | 4,080.79 | 8,110.54 | 2.62 | [-0.24, 0.13] |
| SDMT z-score (Smith et al., 1982) × Stimulation type × Stimulation polarity | 0.04 | 0.01 | 1.00 | 14,179.99 | 20,977.33 | ∞^1^ | [0.03, 0.06] |
| Group × Stimulation type × Stimulation polarity | 0.00 | 0.01 | 1.00 | 15,181.18 | 20,210.41 | 1.54 | [-0.02, 0.02] |
| SDMT z-score (Smith et al., 1982) × Group × Stimulation type × Stimulation polarity | 0.06 | 0.01 | 1.00 | 11,865.37 | 18,779.86 | ∞^1^ | [0.04, 0.08] |
| *Note.* Group-level subject intercepts Estimate: 0.17 (95%CI [0.14, 0.21]). Rhat: 1, Bulk ESS: 5117.68; Tail ESS: 10861.7. Rhat values should be close to 1 to indicate convergence. Bulk ESS and Tail ESS indicate the effective sample size of the MCMC chains. Evidence ratio indicate the ratio of draws that were in the direction of the estimate relative to the number of draws that were in the opposite direction, e.g., for the TMT-A time effect, there were 96.17 times more positive draws than negative draws. Simulations in linear models show, that an evidence ratio of 19 is equivalent to p-value of 0.05 (Makowski et al., 2019). SE=Standard error. ESS=Effective sample size. TMT-A=Trail making test version A. SDMT=Symbol-digits-modalities-test.  ^1^ Evidence ratios equal to ∞ indicate that all posterior draws were in favor of the estimate direction. | | | | | | | |
