## Supplementary material for "Information processing speed modulation by electrical brain stimulation in multiple sclerosis: Towards individually-tailored protocols": Table S10

| **Supplementary Table 11** | | | | | | | |
| --- | --- | --- | --- | --- | --- | --- | --- |
| *Population-level effects of the cumulative probit model for the intensity of adverse effects.* | | | | | | | |
| Parameter | Estimate | *SE* | Rhat | Bulk ESS | Tail ESS | Evidence ratio | 95% CI |
| Intercept_none, mild_ | 1.03 | 0.29 | 1.00 | 2,276.56 | 1,915.80 | 999.00 | [0.44, 1.62] |
| Intercept_mild, moderate_ | 1.95 | 0.30 | 1.00 | 2,349.60 | 2,001.08 | ∞^1^ | [1.32, 2.55] |
| Intercept_moderate, strong_ | 2.85 | 0.34 | 1.00 | 2,726.72 | 1,989.82 | ∞^1^ | [2.18, 3.56] |
| Subject group | -0.14 | 0.19 | 1.00 | 3,202.38 | 3,062.38 | 3.40 | [-0.50, 0.23] |
| Stimulation type | -0.01 | 0.18 | 1.00 | 2,861.39 | 2,863.89 | 1.09 | [-0.37, 0.36] |
| Active stimulation | 0.03 | 0.10 | 1.00 | 8,556.63 | 3,000.71 | 1.64 | [-0.18, 0.23] |
| Itching | 0.21 | 0.39 | 1.00 | 2,607.32 | 2,095.62 | 3.24 | [-0.64, 0.99] |
| Burning | 0.30 | 0.37 | 1.00 | 3,161.52 | 2,491.45 | 4.87 | [-0.48, 1.05] |
| Pain | -0.11 | 0.38 | 1.00 | 2,880.37 | 1,847.70 | 1.80 | [-0.89, 0.72] |
| Metallic taste | -1.66 | 0.59 | 1.00 | 3,926.62 | 2,333.91 | 132.33 | [-2.82, -0.42] |
| Fatigue | 0.37 | 0.39 | 1.00 | 2,493.29 | 2,062.28 | 7.05 | [-0.51, 1.12] |
| Other | 0.37 | 0.38 | 1.00 | 2,217.43 | 1,981.60 | 6.68 | [-0.47, 1.12] |
| Subject group × Stimulation type | 0.11 | 0.26 | 1.00 | 2,743.54 | 3,012.50 | 2.01 | [-0.40, 0.62] |
| *Note.* Group level intercepts *SD_subject_*=0.34, 95%CI=[0.14, 0.54]; *SD_sensation_*=0.27, 95%CI=[0.01, 0.93]. Rhat values should be close to 1 to indicate convergence. Bulk ESS and Tail ESS indicate the effective sample size of the MCMC chains. Evidence ratio indicate the ratio of draws that were in the direction of the estimate relative to the number of draws that were in the opposite direction, e.g., for the TMT-A time effect, there were 239 times more positive draws than negative draws. Simulations in linear models show, that an evidence ratio of 19 is equivalent to a p-value of 0.05 (Markowski et al., 2019).  ^1^ Evidence ratios equal to ∞ indicate that all posterior draws were in favor of the estimate direction. | | | | | | | |
