## Supplementary material for "Information processing speed modulation by electrical brain stimulation in multiple sclerosis: Towards individually-tailored protocols": Table S11

| **Supplementary Table 12** | | | | | | | | | | | | | | | | | | | | | | | |
| --- | --- | --- | --- | --- | --- | --- | --- | --- | --- | --- | --- | --- | --- | --- | --- | --- | --- | --- | --- | --- | --- | --- | --- |
| *Means and standard deviation of the PANAS sum scores for the experimental groups.* | | | | | | | | | | | | | | | | | | | | | | | |
| Valence | Time point | *Control* | | | | | | | | | | *pwRMS* | | | | | | | | | | | |
|  |  | *anodal* | | | | *cathodal* | | | | | | *anodal* | | | | | | *cathodal* | | | | | |
|  |  | *Sham* | | *Active* | | *Sham* | | | *Active* | | | *Sham* | | | *Active* | | | *Sham* | | | *Active* | | |
|  |  | *M* | *SD* | *M* | *SD* | | *M* | *SD* | | *M* | *SD* | | *M* | *SD* | | *M* | *SD* | | *M* | *SD* | | *M* | *SD* |
| positive | pre | 22.00 | 6.00 | 23.38 | 7.54 | | 24.31 | 6.88 | | 23.00 | 8.56 | | 23.29 | 7.01 | | 23.44 | 6.26 | | 22.64 | 6.85 | | 22.64 | 7.27 |
|  | post | 22.38 | 7.93 | 22.94 | 5.79 | | 21.50 | 9.36 | | 21.00 | 8.64 | | 20.79 | 4.59 | | 19.75 | 5.71 | | 19.50 | 8.85 | | 19.50 | 7.06 |
| negative | pre | 1.81 | 2.79 | 1.75 | 4.64 | | 0.44 | 0.51 | | 0.62 | 0.81 | | 1.50 | 1.95 | | 1.69 | 2.02 | | 2.86 | 3.57 | | 2.36 | 2.24 |
|  | post | 0.94 | 1.61 | 0.94 | 3.75 | | 0.00 | 0.00 | | 0.00 | 0.00 | | 1.36 | 1.82 | | 0.69 | 0.70 | | 1.14 | 1.41 | | 1.79 | 2.78 |
| *Note.* M=mean.SD=standard deviation. pwRMS=patients with relapsing multiple sclerosis. | | | | | | | | | | | | | | | | | | | | | | | |
