## Supplementary material for "Information processing speed modulation by electrical brain stimulation in multiple sclerosis: Towards individually-tailored protocols": Table S12

| **Supplementary Table 13** | | | | | | | |
| --- | --- | --- | --- | --- | --- | --- | --- |
| *Fixed effects of the hurdle Gaussian PANAS model.* | | | | | | | |
| Parameter | Estimate | *SE* | Rhat | Bulk ESS | Tail ESS | Evidence ratio | 95% CI |
| Intercept | 20.57 | 0.89 | 1.00 | 8,284.64 | 9,205.50 | ∞^1^ | [18.81, 22.31] |
| Intercept_hurdle_ | -7.53 | 2.19 | 1.00 | 9,271.07 | 5,031.63 | ∞^1^ | [-13.02, -4.63] |
| Valence | -11.36 | 0.75 | 1.00 | 13,662.44 | 9,271.35 | ∞^1^ | [-12.81, -9.87] |
| Time point | 0.26 | 0.65 | 1.00 | 18,761.53 | 9,383.37 | 1.88 | [-1.00, 1.54] |
| Group | 0.09 | 0.79 | 1.00 | 11,748.41 | 10,060.58 | 1.19 | [-1.45, 1.63] |
| Stimulation polarity | 0.41 | 0.79 | 1.00 | 11,325.54 | 9,477.47 | 2.29 | [-1.15, 1.96] |
| Stimulation type | 0.19 | 0.53 | 1.00 | 19,845.19 | 9,228.64 | 1.76 | [-0.86, 1.25] |
| Valence × Time point | -1.40 | 0.81 | 1.00 | 22,119.99 | 8,593.14 | 22.26 | [-2.99, 0.20] |
| Valence × Group | -3.22 | 0.78 | 1.00 | 20,402.31 | 8,781.25 | ∞^1^ | [-4.73, -1.68] |
| Time point × Group | -1.06 | 0.73 | 1.00 | 17,997.89 | 9,377.17 | 12.44 | [-2.49, 0.38] |
| Valence × Stimulation polarity | -2.79 | 0.79 | 1.00 | 19,649.19 | 8,789.27 | 2,999.00 | [-4.34, -1.26] |
| Time point × Stimulation polarity | -0.41 | 0.74 | 1.00 | 19,868.60 | 9,220.28 | 2.47 | [-1.84, 1.02] |
| Group × Stimulation polarity | 0.17 | 0.86 | 1.00 | 13,810.08 | 9,591.41 | 1.37 | [-1.48, 1.87] |
| Time point × Stimulation type | 0.21 | 0.68 | 1.00 | 19,254.29 | 9,412.61 | 1.66 | [-1.11, 1.55] |
| Valence × Time point × Group | -0.39 | 0.88 | 1.00 | 22,328.46 | 8,989.21 | 2.03 | [-2.11, 1.31] |
| Valence × Time point × Stimulation polarity | 0.36 | 0.91 | 1.00 | 21,523.19 | 9,358.34 | 1.90 | [-1.43, 2.15] |
| Valence × Group × Stimulation polarity | -0.26 | 0.85 | 1.00 | 20,364.00 | 9,241.01 | 1.65 | [-1.94, 1.38] |
| Time point × Group × Stimulation polarity | -0.33 | 0.82 | 1.00 | 20,932.56 | 9,891.50 | 1.92 | [-1.95, 1.28] |
| Valence × Time point × Group × Stimulation polarity | 0.35 | 0.93 | 1.00 | 22,480.87 | 9,046.95 | 1.78 | [-1.47, 2.16] |
| Valence_hurdle_ | 7.67 | 2.19 | 1.00 | 9,223.14 | 5,009.22 | ∞^1^ | [4.75, 13.15] |
| *Note.* Subject group-level intercepts Estimate: 4.04 (95%CI [3.21, 4.99]). Rhat: 1, Bulk ESS: 4858.01; Tail ESS7061.43. Rhat values should be close to 1 to indicate convergence. Buld ESS and Tail ESS indicate the effective sample size of the MCMC chains. Evidence ratio indicate the ratio of draws that were in the direction of the estimate relative to the number of draws that were in the opposite direction, e.g., for the Time point effect, there were 1.88 times more positive draws than negative draws. Simulations in linear models show, that an evidence ratio of 19 is equivalent to a p-value of 0.05 (Markowski et al., 2019). SE=Standard error. ESS=Effective sample size.  ^1^ Evidence ratios equal to ∞ indicate that all posterior draws were in favor of the estimate direction. | | | | | | | |
