## Supplementary material for "Information processing speed modulation by electrical brain stimulation in multiple sclerosis: Towards individually-tailored protocols": Table S13

| **Supplementary Table 14** | | | | | | | |
| --- | --- | --- | --- | --- | --- | --- | --- |
| *Fixed effects of the categorical model predicting blinding guess.* | | | | | | | |
| Parameter | Estimate | *SE* | Rhat | Bulk ESS | Tail ESS | Evidence ratio | 95% CI |
| Intercept_t1_ | 0.02 | 0.60 | 1.00 | 13,635.81 | 9,841.71 | 1.06 | [-1.15, 1.18] |
| Intercept_t2_ | 0.53 | 0.54 | 1.00 | 12,311.48 | 10,292.63 | 5.10 | [-0.54, 1.63] |
| Subject group_t1_ | -0.53 | 0.67 | 1.00 | 12,929.49 | 9,602.22 | 3.64 | [-1.85, 0.77] |
| Stimulation type_t1_ | -0.30 | 0.66 | 1.00 | 12,197.62 | 9,508.79 | 2.08 | [-1.61, 0.98] |
| Active stimulation_t1_ | -0.57 | 0.66 | 1.00 | 12,833.65 | 9,288.15 | 4.12 | [-1.87, 0.72] |
| Subject group × stimulation type_t1_ | -0.69 | 0.79 | 1.00 | 13,342.22 | 10,197.73 | 4.33 | [-2.25, 0.84] |
| Subject group × active stimulation_t1_ | -0.69 | 0.79 | 1.00 | 13,809.97 | 9,314.24 | 4.15 | [-2.27, 0.85] |
| Stimulation type × active stimulation_t1_ | 0.91 | 0.75 | 1.00 | 12,100.20 | 10,195.56 | 7.83 | [-0.54, 2.40] |
| Subject group × stimulation type × active stimulation_t1_ | 0.08 | 0.86 | 1.00 | 14,393.74 | 9,470.31 | 1.18 | [-1.63, 1.76] |
| Subject group_t2_ | -0.91 | 0.62 | 1.00 | 11,975.77 | 9,390.54 | 13.32 | [-2.12, 0.28] |
| Stimulation type_t2_ | 0.36 | 0.61 | 1.00 | 12,293.27 | 9,451.60 | 2.61 | [-0.84, 1.56] |
| Active stimulation_t2_ | -0.43 | 0.61 | 1.00 | 12,548.31 | 9,424.39 | 3.11 | [-1.65, 0.79] |
| Subject group × stimulation type_t2_ | -0.42 | 0.74 | 1.00 | 12,905.71 | 9,800.49 | 2.52 | [-1.86, 1.02] |
| Subject group × active stimulation_t2_ | 0.53 | 0.71 | 1.00 | 11,864.54 | 8,666.52 | 3.43 | [-0.87, 1.95] |
| Stimulation type × active stimulation_t2_ | -1.09 | 0.74 | 1.00 | 12,827.28 | 9,196.86 | 13.00 | [-2.52, 0.36] |
| Subject group × stimulation type × active stimulation_t2_ | 0.65 | 0.82 | 1.00 | 12,209.06 | 9,092.66 | 3.77 | [-0.97, 2.25] |
| *Note.* Rhat values should be close to 1 to indicate convergence. Bulk ESS and Tail ESS indicate the effective sample size of the MCMC chains. Evidence ratio indicate the ratio of draws that were in the direction of the estimate relative to the number of draws that were in the opposite direction, e.g., for the TMT-A time effect, there were 239 times more positive draws than negative draws. Simulations in linear models show, that an evidence ratio of 19 is equivalent to a p-value of 0.05 (Makowski et al., 2019). Note that the model fits an every parameter twice: for differences between “do not know”- and “t1”-responses and for differences between “do not know”- and “t2”-responses. | | | | | | | |
