## Supplementary material for "Information processing speed modulation by electrical brain stimulation in multiple sclerosis: Towards individually-tailored protocols": Table S14

**CONSORT: Reporting guidelines checklist of information to include when reporting a randomized trial**

| **SECTION** | **ITEM NUMBER** | **CHECKLIST ITEM** | **REPORTED ON PAGE NUMBER:** |
| --- | --- | --- | --- |
| **TITLE AND ABSTRACT** |  |  |  |
|  | 1a | Identification as a randomized trial in the title |  |
|  | 1b | Structured summary of trial design, methods, results, and conclusions | 2 |
| **INTRODUCTION** |  |  |  |
| Background and objectives | 2a | Scientific background and explanation of rationale | 5 |
|  | 2b | Specific objectives or hypotheses | 6 |
| **METHODS** |  |  |  |
| Trial design | 3a | Description of trial design (such as parallel, factorial) including allocation ratio | 6 |
|  | 3b | Important changes to methods after trial commencement (such as eligibility criteria), with reasons | n/a |
| Participants | 4a | Eligibility criteria for participants | 7 |
|  | 4b | Settings and locations where the data were collected | 7 |
| Interventions | 5 | The interventions for each group with sufficient details to allow replication, including how and when they were actually administered | 8, 9 |
| Outcomes | 6a | Completely defined pre-specified primary and secondary outcome measures, including how and when they were assessed | 10 |
|  | 6b | Any changes to trial outcomes after the trial commenced, with reasons | n/a |
| Sample size | 7a | How sample size was determined | 7 |
|  | 7b | When applicable, explanation of any interim analyses and stopping guidelines | n/a |
| Randomisation: |  |  |  |
| Sequence generation | 8a | Method used to generate the random allocation sequence | 7 |
|  | 8b | Type of randomisation; details of any restriction (such as blocking and block size) | 7 |
| Allocation concealment mechanism | 9 | Mechanism used to implement the random allocation sequence (such as sequentially numbered containers), describing any steps taken to conceal the sequence until interventions were assigned | 7 |
| Implementation | 10 | Who generated the random allocation sequence, who enrolled participants, and who assigned participants to interventions | 7 |
| Blinding | 11a | If done, who was blinded after assignment to interventions (for example, participants, care providers, those assessing outcomes) and how | 9 |
|  | 11b | If relevant, description of the similarity of interventions | 9 |
| Statistical methods | 12a | Statistical methods used to compare groups for primary and secondary outcomes | 10-14 |
|  | 12b | Methods for additional analyses, such as subgroup analyses and adjusted analyses | 11, 12 |
| **RESULTS** |  |  |  |
| Participant flow (a diagram is strongly recommended) | 13a | For each group, the numbers of participants who were randomly assigned, received intended treatment, and were analyzed for the primary outcome | 7, 8 |
|  | 13b | For each group, losses and exclusions after randomisation, together with reasons | 8 |
| Recruitment | 14a | Dates defining the periods of recruitment and follow-up | n/a |
|  | 14b | Why the trial ended or was stopped | n/a |
| Baseline data | 15 | A table showing baseline demographic and clinical characteristics for each group | Table 1 |
| Numbers analyzed | 16 | For each group, number of participants (denominator) included in each analysis and whether the analysis was by original assigned groups | 7, 8, Table 1 |
| Outcomes and estimation | 17a | For each primary and secondary outcome, results for each group, and the estimated effect size and its precision (such as 95% confidence interval) | 15, 16, 17 |
|  | 17b | For binary outcomes, presentation of both absolute and relative effect sizes is recommended | n/a |
| Ancillary analyses | 18 | Results of any other analyses performed, including subgroup analyses and adjusted analyses, distinguishing pre-specified from exploratory | 16 |
| Harms | 19 | Important harms or unintended effects in each group | 17, 18, 19 |
| **DISCUSSION** |  |  |  |
| Limitations | 20 | Trial limitations, addressing sources of potential bias, imprecision, and, if relevant, multiplicity of analyses | 23 |
| Generalizability | 21 | Generalizability (external validity, applicability) of the trial findings | 23 |
| Interpretation | 22 | Interpretation consistent with results, balancing benefits and harms, and considering other relevant evidence | 19-22 |
| **Other information** |  |  |  |
| Registration | 23 | Registration number and name of trial registry | 6 |
| Protocol | 24 | Where the full trial protocol can be accessed, if available | n/a |
| Funding | 25 | Sources of funding and other support (such as supply of drugs), role of funders | 26 |

If relevant, CONSORT extensions for cluster randomized trials, non-inferiority and equivalence trials, non-pharmacological treatments, herbal interventions and pragmatic trials are available www.consort-statement.org
