## Supplementary material for "Information processing speed modulation by electrical brain stimulation in multiple sclerosis: Towards individually-tailored protocols": Table S15

| **Supplementary Table 1** | | | | | | | | |
| --- | --- | --- | --- | --- | --- | --- | --- | --- |
| *Absolute number and percentage (relative to all trials after each filtering step) of included trials.* | | | | | | | | |
|  | *Control* | | | | *MS Patients* | | | |
|  | *anodal* | | *cathodal* | | *anodal* | | *cathodal* | |
|  | *Sham* | *Active* | *Sham* | *Active* | *Sham* | *Active* | *Sham* | *Active* |
| All trials | 7652 (100%) | 7878 (100%) | 8789 (100%) | 8536 (100%) | 5918 (100%) | 5785 (100%) | 5685 (100%) | 6021 (100%) |
| Step (1) | 7543 (98.58%) | 7743 (98.29%) | 8631 (98.2%) | 8370 (98.06%) | 5776 (97.6%) | 5672 (98.05%) | 5592 (98.36%) | 5937 (98.6%) |
| Step (2) | 7541 (98.55%) | 7739 (98.24%) | 8628 (98.17%) | 8361 (97.95%) | 5757 (97.28%) | 5652 (97.7%) | 5566 (97.91%) | 5924 (98.39%) |
| Step (3) | 7220 (94.35%) | 7410 (94.06%) | 8203 (93.33%) | 7972 (93.39%) | 5500 (92.94%) | 5437 (93.98%) | 5344 (94%) | 5678 (94.3%) |
| *Note.* Data were cleaned in the following order: (1) Removal of incorrect responses, (2) Removal of trials with response times outside the 0.2-6s interval, (3) Removal of trials with response latencies outside of the individual median ± 3 × MAD interval. The individual median was calculated for each subject and session. MAD=Median Absolute Deviation. s=seconds. | | | | | | | | |
